# The Contribution of Common and Rare Genetic Variation to Emotional and Behavioural Symptoms in Childhood and Adolescence

**DOI:** 10.1101/2025.07.01.25330628

**Authors:** Olivia Wootton, Emma E. Wade, Daniel S. Malawsky, Mahmoud Koko, Qin Qin Huang, Varun Warrier, Matthew E. Hurles, Hilary C. Martin

## Abstract

Genetic factors influence vulnerability to common mental health conditions, but their role in early-life mental health remains understudied. We analysed genotype array and exome sequence data from two birth cohorts (Millenium Cohort Study and Avon Longitudinal Study of Parents and Children; n=5,320-8,622) to assess common and rare variant contributions to internalising and externalising symptoms across development. For both symptom domains, we identified associations with several polygenic indices (PGIs) that generally remained stable across development. For externalising symptoms, many of these associations reflected direct genetic effects. Concordant results were observed in the Born in Bradford cohort. A higher exome-wide burden of deleterious rare variants was associated with increased externalising (p-adj<0.03) and internalising symptoms (p-adj<0.01); trio models indicated direct genetic effects on externalising in MCS (p<0.05; p-adj>0.05) and on internalising symptoms in ALSPAC (p-adj<0.02). Common and rare genetic variants contributed independently, jointly explaining 2% of the variance in internalising and 5-7% in externalising symptoms. Finally, we show that the direct genetic effects of several PGIs and of deleterious rare variants on adolescent mental health are mediated by childhood externalising behaviours and/or cognitive ability. This study provides new insights into the genetic architecture of early-life mental health and identifies promising avenues for future research.

## Introduction

Mental health conditions that emerge in childhood and adolescence often persist into adulthood^1^ and are associated with a range of adverse health and developmental outcomes.^2–9^ Both genetic and environmental factors contribute to early-life emotional and behavioural difficulties.^10^ Twin studies estimate the heritability of internalising disorders (anxiety and depressive disorders) to be 30-50%,^11,12^ whereas externalising disorders (Attention Deficit Hyperactivity Disorder (ADHD), conduct disorder, and oppositional defiant disorder) have higher heritability estimates of 50-80%^10,13^.

Understanding how genetic variation influences mental health traits across development and how these influences evolve over time may provide a clearer picture of behavioural trajectories. Genetic variants carried by a child may influence their phenotype directly (direct genetic effects); additionally, as children share genetic variants with their parents, these effects may be indirect and act via genetically-influenced parental characteristics (indirect genetic effects).^14^ By adjusting for parental genotypes in tests for an association between a child’s genotype and a trait, it is possible to estimate direct genetic effects as well as the effects of non-transmitted alleles, which capture indirect genetic effects and confounders like assortative mating and population stratification.^14–17^

Studies investigating direct and indirect genetic effects on early emotional and behavioural traits have so far focused on common variation, and yielded mixed results, likely due to methodological heterogeneity and the limited availability of large, well-powered family-based cohorts. For externalising traits (e.g., impulsivity, hyperactivity, and aggression), some studies have found evidence for only direct genetic effects^18–20^, while others have identified both direct effects and effects of non-transmitted parental alleles^21–26^. Two recent studies that reported a direct genetic effect of polygenic predisposition to adult externalising behaviours on externalising symptoms found that the magnitude of the association varied across development - peaking during childhood in one study^20^, and increasing from childhood to adolescence in the other^24^. Research on internalising traits (eg. anxiety and depression) remains limited. Although there is evidence for both direct and indirect genetic associations with childhood internalising difficulties, existing research is based on data from the same cohort^22,25,27^ and to our knowledge, only one study considers changes over time ^28^. Both replication in independent samples and additional longitudinal analyses are needed to clarify these associations. Finally, recruitment into family-based genetic studies is frequently subject to participation biases^17,29^ - an issue that extends to work on mental health symptoms - potentially limiting the generalisability of findings and introducing biases into genetic effect estimates^30,31^.

Common variants account for less than half of the total heritability estimated from twin studies for early emotional and behavioural traits.^32–34^ It is unknown how much of this “missing heritability” is attributable to rare variants. Rare copy number variants (CNVs) associated with developmental disorders are known to influence childhood mental health in both population-based^35^ and clinically-ascertained cohorts^36–38^. Deleterious sequence-level mutations, such as protein-truncating and damaging missense variants, also contribute to related traits, like cognitive ability and educational attainment ^39,40^, and to mental health conditions in adulthood.^39,41,42^.

It is also important to understand whether and how genetic influences on emotional and behavioural problems are mediated by intermediate traits. Previous research suggests that childhood externalising behaviours are correlated with risk for both externalising and internalising disorders in adolescence^43,44^, and that childhood cognitive ability and related constructs, e.g. academic performance, are associated with later mental health outcomes^45–49^. Thus, childhood externalising behaviours and cognitive ability pose two potential pathways through which genetic variation may influence adolescent mental health. Supporting this, studies using genetically-informed designs have demonstrated a causal relationship between genetic predisposition to ADHD and major depressive disorder.^50,51^ However, it remains unclear whether genetic predisposition to ADHD and other traits affects adolescent mental health directly or through mediation by childhood traits. Furthermore, to our knowledge, previous studies have not explored mediators of genetic factors using direct genetic effect estimates, which are more robust to bias from confounders like population stratification and assortative mating^52–54^.

In this work, we leverage genotype array and new exome-sequence data from three birth cohorts to investigate genetic risk factors for childhood and adolescent mental health symptoms. First, we use mixed effects modelling with several measures of common and rare genetic variation to evaluate associations with repeated measures of internalising and externalising symptoms. We estimate these associations using the full ‘population’ sample and within families (i.e. adjusting for parental genetic scores) to evaluate the contribution of direct genetic effects, and test for changes in these associations over time. Next, we investigate whether common and rare genetic variation contribute independently to variation in internalising and externalising symptoms and quantify the relative importance of each genetic measure for prediction of these traits. Finally, we examine whether childhood traits mediate direct genetic effects on adolescent internalising and externalising problems. By integrating common and rare variant analyses with within-family designs, we aim to refine understanding of the mechanisms underlying these traits and inform future research into early identification and prevention of mental disorders.

## Results

We analysed genotype array and exome-sequence data from three British birth cohorts that recruited participants across three decades: the Avon Longitudinal Study of Parents and Children (ALSPAC)^55,56^, the Millennium Cohort Study (MCS)^57^, and Born in Bradford (BiB)^58,59^ (**Figure 1A and 1B**). Participants were recruited from different regions of the UK and demographic composition differed between studies. ALSPAC participants were recruited between 1991 and 1992 from the Bristol area of southwest England and are typically from advantaged socioeconomic backgrounds; MCS was a nation-wide cohort recruited between 2000 and 2002 that aimed to be broadly representative while deliberately oversampling participants from disadvantaged and ethnic minority backgrounds; and BiB participants were recruited between 2007 and 2011 from an ethnically diverse population in Bradford in the north of England, with the majority from low socioeconomic status households. The sex distribution was comparable across all cohorts, with males comprising 48-51% of the participants.

**Figure 1.**
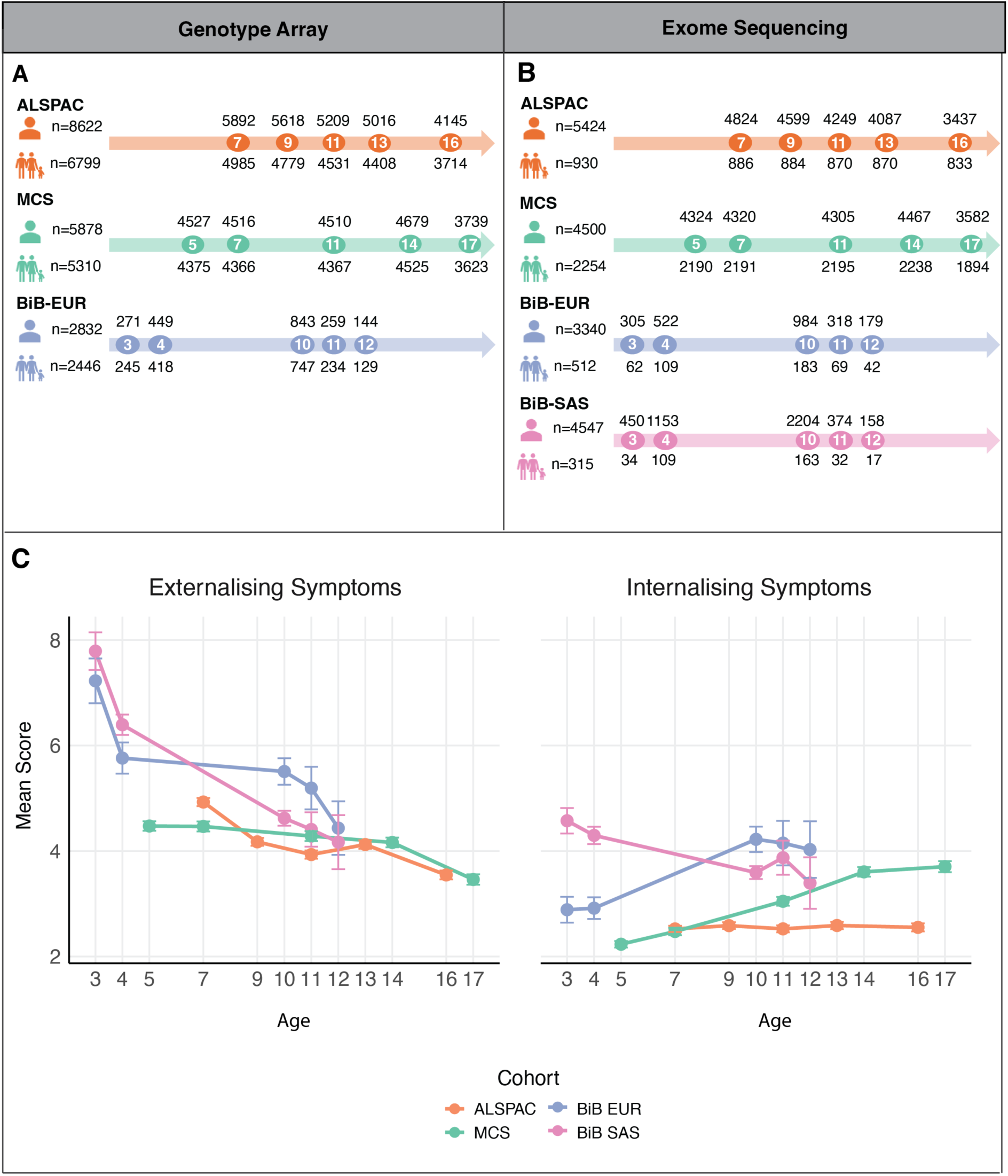
Overview of the birth cohort data. **A–B**) For each cohort, the number of participants with genotype array (**A**) and exome sequence data (**B**) is shown to the left of the arrow. Each assessment age is represented by an opaque circle with the number of children with Strengths and Difficulties Questionnaire (SDQ) and genetic data above the arrows, and the number of children in parent–child trios shown below. Note that, for genotype array data, the latter includes imputed trios following Mendelian imputation of missing parental genotypes for children genotyped as duos. **C**) Mean externalising and internalising SDQ subscale scores at each assessment age for children with genetic data. Error bars represent 95% confidence intervals.

In each study, internalising and externalising symptoms were assessed using the Strengths and Difficulties Questionnaire (SDQ), a well-validated 25-item questionnaire used to screen for mental health problems in community samples of children and adolescents.^60^ The SDQ was completed by parents at five timepoints spanning childhood and adolescence. On average, externalising symptoms were highest at the earliest assessment age across all studies (**Figure 1C**; **Supplementary Figure 1**). Internalising symptom scores remained relatively stable over time, except in MCS, in which scores increased with age. For both symptom domains, mean scores were higher among BiB participants compared to those in MCS and ALSPAC. With the exception of age seven, symptom scores in MCS were higher than in ALSPAC or did not differ significantly. We used data from ALSPAC and MCS for the main analyses because of their larger sample sizes and similarities in the distribution of SDQ symptom scores and the range of assessment ages; BiB analyses are reported separately due to differences in these characteristics (**Figure 1**; **Supplementary Figure 1**).

### Influence of common variants on internalising and externalising symptoms across development

We first examined the associations between internalising and externalising symptoms and common variant predisposition, measured using polygenic indices (PGIs), to conditions on the internalising and externalising spectrum, and to education-related traits. We fitted linear mixed-effects models in which symptom scores across ages were regressed on individual-specific random intercepts and fixed effects, including the PGI, age and sex (<u>Methods</u>). In these models, the main genetic effect (referred to as the “population effect”) represents the association between the PGI and symptom score at the earliest assessment age, estimated by leveraging data across all ages. Across both ALSPAC and MCS (N_ALSPAC_=6,690; N_MCS_=4,709), higher externalising symptoms were significantly associated with higher polygenic predisposition to externalising behaviours, ADHD, neuroticism, and depression, and with lower predisposition to educational attainment (EA) and its cognitive and non-cognitive components (Benjamini-Yekutieli adjusted p(p-adj)<1.1x10^-3^) (**Figure 2**; **Supplementary Table 1**). Higher internalising symptoms were significantly (p-adj<6.3x10^-3^) associated with increased polygenic predisposition to ADHD, neuroticism, generalised anxiety disorder, and depression.

**Figure 2.**
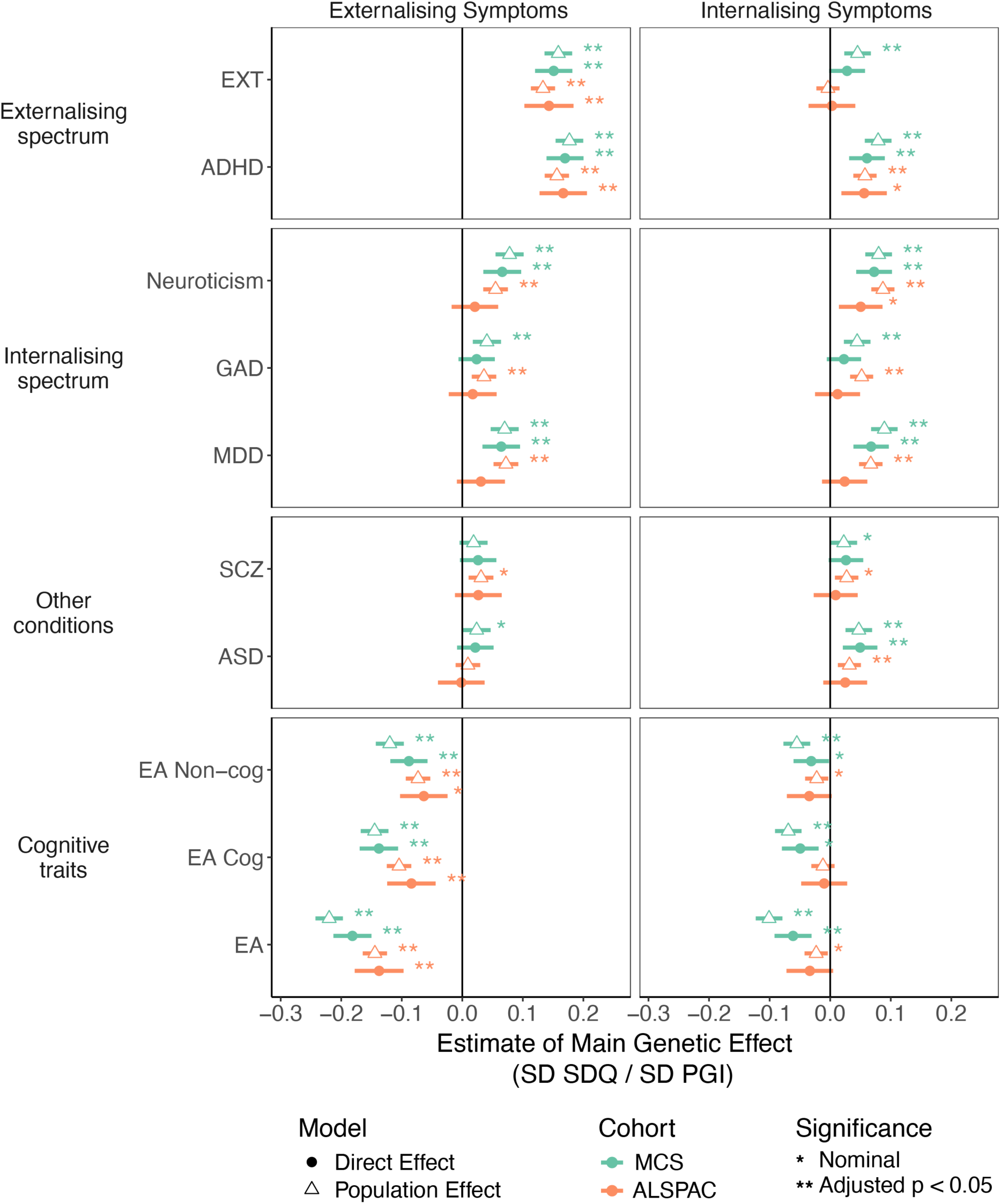
Associations between polygenic indices (PGIs) and childhood and adolescent externalising and internalising symptoms. Standardised effects and 95% confidence intervals estimated for the main effects of the genetic score per standard deviation change in symptom score. Population effect sizes were estimated in the full sample (triangle) and direct genetic effects were estimated in models controlling for parental genetic scores (shaded circles). ADHD: Attention-deficit hyperactivity disorder; ASD: Autism spectrum disorder; EA: educational attainment; EA Cog: cognitive component of EA; EA Non-cog: non-cognitive component of EA; EXT: externalising behaviours; GAD: Generalised anxiety disorder; MDD: Major depressive disorder; SCZ: schizophrenia.

We then sought to determine whether the observed associations between a child’s genotype and their symptoms reflects direct genetic effects, or the influence of factors correlated with the child’s genotype, such as genetically-influenced parental behaviours and confounders. To estimate direct genetic effects, we jointly fitted the child and parental PGIs in the linear mixed-effects model (“trio model”). As many children only had a single parent genotyped in the study, we used Mendelian imputation to impute the missing parental genotypes. This increased the sample size of genotyped trios from 1,471 to 6,799 in ALSPAC and from 2,224 to 5,310 in MCS. Before running the trio model, we confirmed that the results above (i.e., the PGI association with each symptom domain) were consistent whether analysing probands from the full cohort (Population Effect - Whole Cohort) or the subset of probands with at least one parent genotyped (Population Effect - Trios) (**Supplementary Figure 2**; **Supplementary Table 1**).

Our findings from the trio model suggest that the associations between externalising symptoms and polygenic predisposition to externalising behaviours and ADHD are explained by direct genetic effects. However, the association with PGIs for EA and its non-cognitive component (EA non-cog) may be due to direct genetic effects as well as genetically-influenced maternal traits and behaviours, or potential confounding. Specifically, we found at least nominally significant direct effects of PGIs for externalising behaviours, ADHD, and education-related traits on externalising symptoms across both cohorts (p<0.05; p-adj>0.05) (**Figure 2**; **Supplementary Table 1**), while we only saw significant non-transmitted coefficients for the EA and EA non-cog PGIs, and then only for mothers in MCS (p-adj<0.04) (**Supplementary Figure 3**; **Supplementary Table 2**). For internalising symptoms, in both cohorts, we observed at least nominally significant direct effects for the ADHD and neuroticism PGIs, as well as associations with the maternal PGIs for neuroticism, anxiety, and depression (**Figure 2**; **Supplementary Figure 3**).

We observed concordant results for population estimates using SDQ symptoms measured at a single time point for BiB participants (<u>Methods</u>; **Supplementary Figure 4**; **Supplementary Table 3**). For estimates from the trio models, the direction of direct genetic effect was generally consistent with that observed in MCS and ALSPAC, but confidence intervals were wide due to the modest sample size (N=1,037).

Returning to ALSPAC and MCS, we also fitted models with an interaction between age and each PGI to test whether these genetic associations changed across development (<u>Methods</u>). Across externalising and internalising symptoms, only the externalising behaviour PGI showed evidence of an interaction with age in the population models, suggesting that the influence of common variants associated with this trait on externalising symptoms increased with age (p<0.05; p-adj>0.05) (**Supplementary Figure 5**; **Supplementary Table 4**). Moreover, the increasing influence of these variants with age appears to be driven by direct genetic effects: we observed a nominally significant positive interaction between the direct effect of the externalising behaviour PGI and externalising symptoms in the trio models in both ALSPAC and MCS (p<0.05; p-adj>0.05) (**Supplementary Figure 5**; **Supplementary Table 4**).

### Influence of rare variant burden on internalising and externalising symptoms

Next, we tested whether rare genetic variation influences childhood mental health using rare variant burden scores (RVBS). These scores were calculated by summing gene-specific selection coefficients^61^ for genes in which an individual carries a rare predicted loss-of-function protein truncating variant (RVBS_PTV_) or damaging missense variant (RVBS_missense_). In linear mixed effects models considering all children, we observed a significant main effect of RVBS_PTV_; higher RVBS_PTV_ was associated with an increase in externalising (p-adj<0.03) and internalising (p-adj<0.01) symptoms in population models in both cohorts (N_ALSPAC_=5,424; N_MCS_=4,500) (**Figure 3**; **Supplementary Table 5**). RVBS_missense_ showed a nominally significant (p<0.05; p-adj>0.05) association with increased internalising symptoms in both cohorts, and with increased externalising symptoms in MCS. Direct effect estimates from trio models were consistent across cohorts for externalising (*Z*=0.97, p=0.33) but not internalising (*Z*=-2.7, p=6.3x10^-3^) symptoms. RVBS_PTV_ showed a significant direct effect on internalising symptoms in ALSPAC (p-adj<0.02) but not in MCS. No significant associations were observed for parental non-transmitted coefficients in either cohort (**Supplementary Figure 6**).

**Figure 3.**
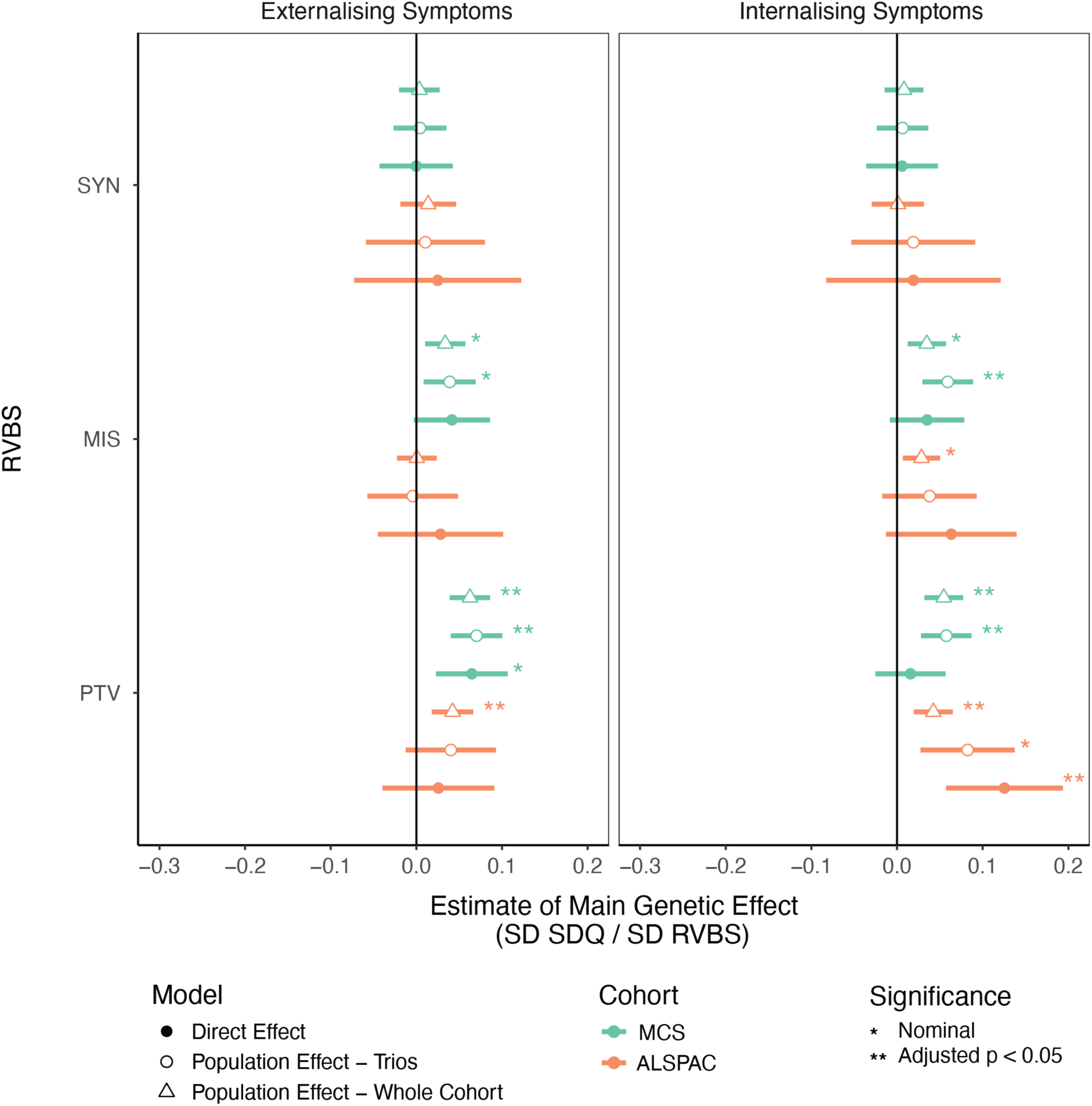
Associations between rare variant burden scores (RVBS) and childhood and adolescent externalising and internalising symptoms. Standardised effects (in standard deviations of SDQ score) and 95% confidence intervals estimated for the main effects of the RVBS per standard deviation change in symptom score for RVBS calculated with three different consequence classes. Population effect sizes were estimated in the full sample (circle) and in a subset of children with parental RVBS (triangle). Shaded circles illustrate direct effect estimates from models controlling for parental genetic scores. MIS: missense; PTV: protein truncating variant; SYN: synonymous.

Finally, we estimated associations between RVBS and symptoms separately in the two main genetically-inferred ancestry groups from BiB: EUR (N=1,632; N_trios_=292) and SAS (N=3,118; N_trios_=224). For the EUR group but not the SAS group, we replicated the positive association between RVBS_PTV_ and both symptom domains in the population models including all children, at a nominal level of significance (p<0.05; p-adj>0.05) (**Supplementary Figure 7**; **Supplementary Table 6**). Despite differences in statistical significance, formal tests for heterogeneity did not indicate meaningful differences in effect sizes between the ancestry groups (Bonferroni corrected Z-test p-value < 0.05; **Supplementary Table 7**). No significant direct effects were observed for either genetic ancestry group, likely due to limited power.

Across all models and cohorts, the association with RVBS for synonymous variants was null, as expected, indicating that the observed associations with PTV and missense RVBS are unlikely to be fully explained by uncontrolled population stratification. Taken together, these results show that burden of deleterious PTVs is associated with both internalising and externalising symptoms in childhood and adolescence.

### Relative contribution of common and rare genetic variation to internalising and externalising symptoms

Our findings so far demonstrate a relationship between childhood mental health and both common and rare genetic variation. To explore this further, we used a stepwise regression approach to assess the relative contribution of each genetic measure to variance in internalising and externalising symptoms. We started by ranking the genetic measures based on the variance they explained in each symptom domain (marginal *R*^2^) (**Supplementary Table 8**) and then added them sequentially to a conditional model, retaining a measure only if it significantly improved model fit (log-likelihood ratio test adjusted p-value < 0.05). In the best fitting conditional models, genetic measures explained more variance in externalising symptoms (4-7%) than internalising symptoms (∼2%) (**Supplementary Table 9**). Measures of rare genetic variation contributed more to the prediction of internalising symptoms than externalising symptoms; specifically, adding them to the baseline model increased the variance explained by the fixed effects by 10-13% for internalising symptoms and by 2-4% for externalising symptoms, although the overall variance explained by the rare variant burden scores was low (0.1-0.3%) (**Supplementary Table 10**).

Next, we assessed whether the associations between genetic measures and symptoms reflected independent contributions or shared genetic liability across predictors by comparing effect size estimates from conditional and marginal models. Marginal models included a single genetic measure with covariates, while conditional models included multiple genetic measures. We observed an attenuation in the effect sizes of the PGIs in conditional compared to marginal models (**Figure 4**; **Supplementary Table 11**), which may reflect a shared genetic liability across these traits, consistent with the concept of a general psychopathology (p) factor^62,63^. However, for both symptom domains, the effect size estimate for the measures of rare genetic variation remained largely unchanged in the conditional model. These results indicate that the contributions of individual PGIs to externalising and internalising symptoms are not entirely independent when other PGIs are considered. In contrast, the influence of rare genetic variation, indexed by RVBS_PTV_ and RVBS_MIS_, appears to be distinct from that of common genetic factors.

**Figure 4.**
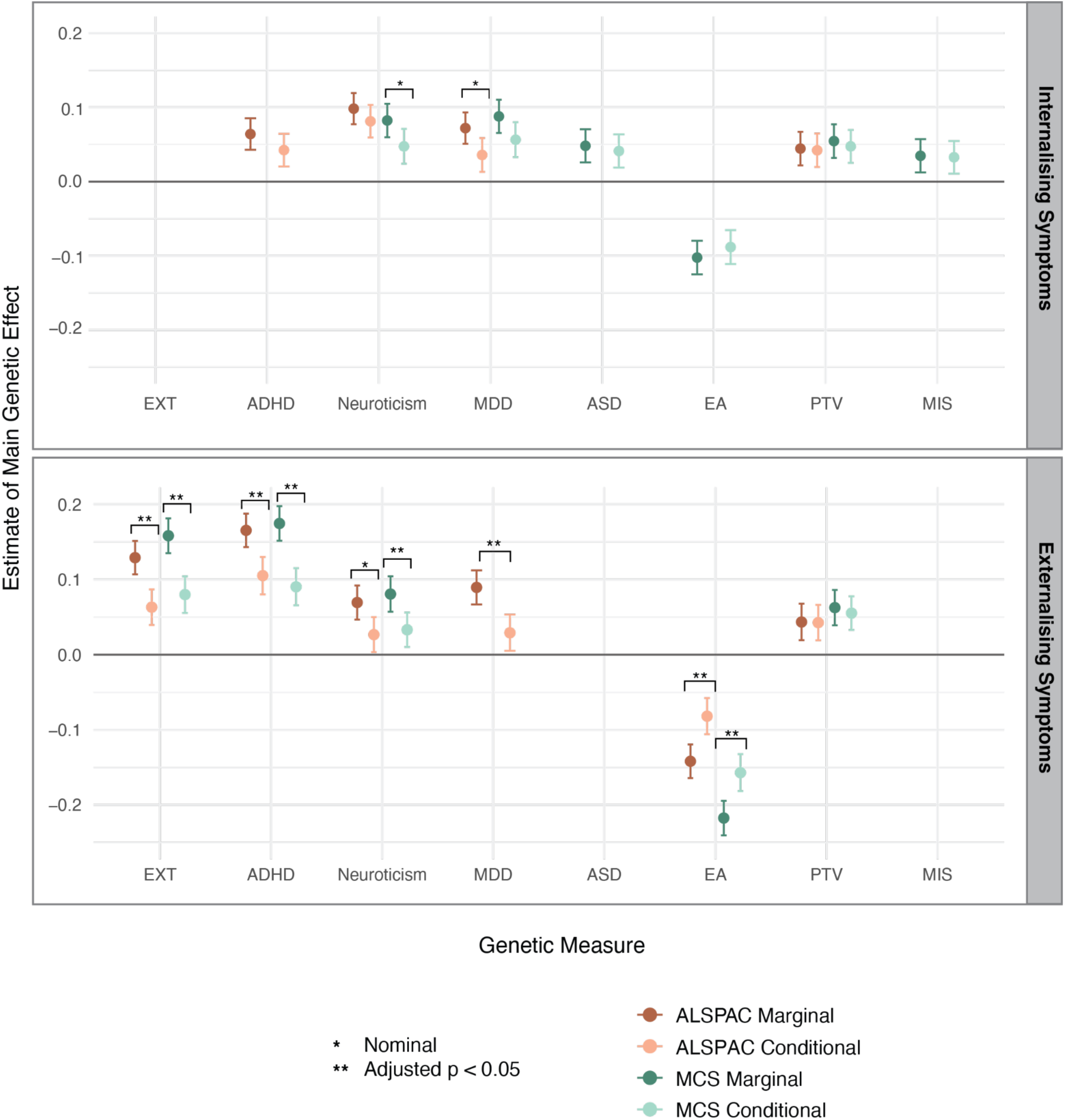
Associations between genetic measures and internalising and externalising symptoms. Standardised effect sizes and 95% confidence intervals are shown for the main effects of each genetic measure, estimated using two models per cohort: (1) marginal models including only the indicated genetic measure and standard covariates, and (2) conditional models including all genetic measures as predictors. Analysis was limited to genetic measures that showed at least a nominally significant association with the symptom domain in the population effect models. Asterisks indicate significant differences between marginal and conditional estimates based on Z-tests. ADHD: Attention-deficit hyperactivity disorder; ASD: Autism spectrum disorder; EA: educational attainment; EXT: externalising behaviours; MIS: missense; MDD: Major depressive disorder; PTV: protein truncating variant.

### Mediators of genetic influences on adolescent internalising and externalising symptoms

Finally, we sought to investigate the mechanisms through which genetic variation influences mental health across development. Evidence suggests that certain childhood traits and behaviours, such as externalising difficulties and cognitive ability, are associated with mental health conditions in adolescence.^43–48^ We conducted mediation analyses to explore whether genetic associations with adolescent mental health symptoms may be explained, in part, by earlier-emerging traits (**Figure 5A**). In this context, we define the *mediated path* as the portion of the genetic association with adolescent mental health symptoms that operates through the specified childhood trait, and the *unmediated path* as the portion that remains after accounting for the association with the mediator. We note that associations acting via the “unmediated” path in this framework may still be mediated by other unmeasured intermediate traits not included in our models and therefore should not be interpreted as strictly unmediated. We focused on estimating mediation of direct genetic effects, and thus only considered genetic measures with at least a nominally significant direct effect on both the mediator (the childhood trait) and the outcome (adolescent internalising or externalising symptoms) (**Supplementary Figure 8-11**).

**Figure 5.**
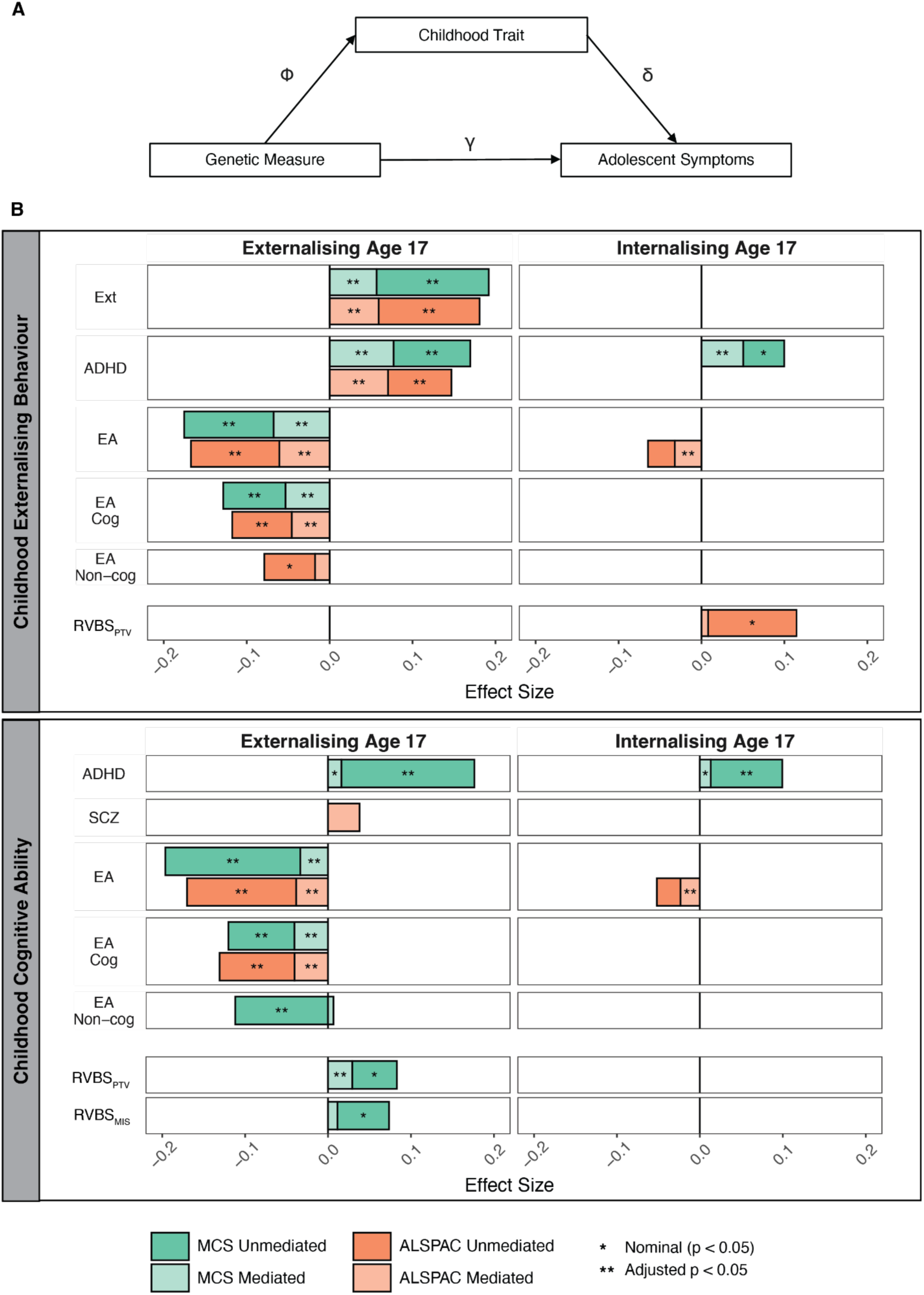
Mediation of direct genetic effects on adolescent externalising and internalising via childhood traits. **A)** Path diagram illustrating the decomposition of the direct genetic effect of a genetic measure on adolescent mental health symptoms into two parts - the unmediated pathway (γ) and the mediated pathway acting via a childhood trait (the product of Φ and δ). **B**) Stacked bar chart illustrating the effect size of a genetic measure on adolescent externalising and internalising symptoms mediated by childhood externalising behaviour (top panel) and childhood cognitive ability (bottom panel). For each genetic measure, bars represent standardised effect sizes from the unmediated path (γ) and the mediated path (Φ × δ). Bars are displayed only for genetic measures that showed an association with both the mediator and the outcome within a given cohort. ADHD: Attention-deficit hyperactivity disorder; EA: educational attainment; EA Cog: cognitive component of EA; EA Non-cog: non-cognitive component of EA; Ext: externalising behaviours; MIS: missense; PGI: polygenic index; PTV: protein truncating variant; RVBS: rare variant burden score.

We first considered whether externalising behaviours in childhood mediated genetic associations with adolescent externalising and internalising symptoms. All but one of the tested PGIs (externalising behaviours, ADHD, EA and its cognitive component) influenced adolescent externalising symptoms via both the unmediated (p-adj<0.03) and mediated path (p-adj<4.6x10^-3^) (**Figure 5B**, top left; **Supplementary Table 12**). The exception was the EA non-cog PGI, for which we found no evidence of mediation via childhood externalising behaviours. For adolescent internalising symptoms, the direct genetic effects of PGIs for EA and ADHD were mediated via their association with childhood externalising behaviours (p-adj<2.2x10^-4^), whereas RVBS_PTV_ acted via the unmediated path (p<0.05; p-adj>0.05) (**Figure 5B**, top right; **Supplementary Table 12**).

We then considered whether childhood cognitive ability mediated genetic associations with adolescent externalising and internalising symptoms. We also found that the associations between adolescent externalising symptoms and the PGIs for EA and its cognitive component, as well as RVBS_PTV_, were partly mediated via childhood cognitive ability (p-adj<2.3x10^-4^) (**Figure 5B**, bottom left; **Supplementary Table 13**). In contrast, the PGI for the non-cognitive component of EA influenced adolescent externalising symptoms exclusively via the unmediated pathway (p-adj<2.3x10^-4^). The association between the EA PGI and adolescent internalising symptoms was fully mediated via childhood cognitive ability (p-adj<5.3x10^-4^) (**Figure 5B**, bottom right; **Supplementary Table 13**). Taken together, these findings suggest that several genetic influences on adolescent mental health act through both unmediated pathways as well as via pathways mediated by their association with childhood traits.

## Discussion

This study provides new evidence for the role of both common and rare genetic variants in shaping internalising and externalising symptoms across childhood and adolescence. Key strengths include: 1) leveraging longitudinal phenotypic data through linear mixed-effects models to account for intra-individual correlations and increase power for genetic association analyses, 2) imputing missing parental genotypes to increase sample size and reduce bias in our common variant analyses, and 3) jointly modelling common and rare genetic variation to address existing gaps in understanding how genetic variation across the allele frequency spectrum influences early-life mental health.

In our analysis of common genetic variation, we found that associations between externalising symptoms and PGIs for externalising behaviours, ADHD, and the cognitive component of EA were primarily explained by direct genetic effects (**Figure 2**). These associations were generally consistent across cohorts and correspond with previous work^18–20^, though findings have varied across studies^21–23,25,26^. We found limited evidence for associations between non-transmitted parental alleles and externalising symptoms (**Supplementary Figure 3**). An exception was the negative association between externalising symptoms and the maternal PGIs for EA and its non-cognitive component in MCS (**Supplementary Figure 3**). These results indicate that the significant population effects for these PGIs may also include associations with genetically-influenced maternal characteristics, such as parenting behaviours or the prenatal environment, or unmeasured confounding^14–17^. An association between the maternal non-transmitted coefficient for EA and offspring externalising traits has also been observed in MoBA, a Norwegian birth cohort, although the direction of effect differed from our findings.^18,64^ Our results align more closely with the epidemiological observation that lower household socioeconomic status, including lower parental EA, is associated with increased externalising problems in children.^65,66^ Given these findings, clarifying the mechanisms underlying intergenerational transmission of risk, whether through true indirect genetic effects or residual confounding, should be a focus of future research.

For internalising symptoms, we found evidence of a positive direct genetic effect of PGIs for ADHD and neuroticism, reaching at least nominal significance in both cohorts (**Figure 2**). The nominally significant positive association between internalising symptoms and maternal PGIs for neuroticism, depression, and anxiety may reflect indirect genetic effects or confounding factors, including the possibility that maternal genetic predisposition to these traits influences reporting on their child’s internalising symptoms^67,68^, and warrants further investigation (**Supplementary Figure 3**). More broadly, the paucity of associations passing multiple testing correction in the trio models may reflect the lower heritability of internalising traits^32^, which would reduce power to detect associations.

One methodological advantage of this study is the use of Mendelian imputation to recover missing parental genotypes, whereas previous studies have typically relied on complete genotyped trios^18,20,21,23^. By using Mendelian imputation, we substantially increased the number of genotyped trios - from 1,471 to 6,799 in ALSPAC and from 2,224 to 5,310 in MCS - improving power and increasing the significance of several PGI associations. This approach also yielded more consistent findings across cohorts (**Extended Data Figure 1; Supplementary Figure 12 ; Supplementary Note 1**) and, in some cases, led to more robust conclusions than analyses restricted to complete trios (**Extended Data Figure 2**; **Supplementary Figure 13**). For example, in ALSPAC, the associations between externalising symptoms and the direct genetic effects of PGIs for externalising behaviours and EA were non-significant when using only observed trios but became significant and increased in magnitude when using imputed trios. Previous research has found evidence of education-related ascertainment bias in these cohorts, showing that children in fully genotyped trios have a higher EA PGI than children who do not have genetic data on both parents.^17^ Thus, the changes in our results when using imputed rather than complete genotyped trios likely reflect not only increased sample size but also a reduction in bias introduced by non-random trio ascertainment.

Additionally, the inclusion of rare exonic variation directly addresses a substantial limitation of existing studies, which have predominantly focused on common genetic variants. We demonstrate that a higher burden of deleterious PTVs is associated with increased internalising and externalising symptoms (**Figure 3**). Notably, we show that RVBS_PTV_ explains additional variance in both symptom domains beyond that accounted for by common genetic variation in conditional models (**Figure 4**). These findings support the hypothesis that some of the “missing heritability” (i.e., the difference between twin-based and SNP-based heritability estimates) may be attributable to the effects of rare variants, and imply that combining common and rare variant measures could improve prediction of early mental health outcomes. Whole-genome sequence data in very large samples will be required to quantify the total contribution of rare variation (including noncoding variants) to these traits ^69^.

We conducted mediation analysis to better understand the pathways through which genetic variation influences adolescent mental health symptoms. Our results show that both externalising behaviours and cognitive ability in childhood partially mediate the direct genetic effects of multiple PGIs on adolescent symptoms (**Figure 5**). These findings are consistent with previous research^43–49^ and with the developmental cascades theory, which posits that early-life traits or behaviours can trigger a chain of interrelated processes that shape later outcomes.^70^ Importantly, by controlling for parental PGIs in our mediation models, we reduce bias from indirect genetic effects and other confounders^14–17^, providing a more robust basis for identifying pathways through which direct genetic liability may influence adolescent outcomes. Taken together, our results indicate that while genetic predisposition to some traits influences adolescent mental health symptoms directly, part of this association is mediated via childhood externalising behaviours and cognitive ability. This suggests that, in certain cases, childhood may represent a meaningful window for modifying the impact of genetic factors on adolescent mental health.

Our study has several limitations. First, using SDQ symptom scores from a single rater may have introduced reporter bias, as cross-informant agreement on SDQ is typically low to moderate in population-based samples.^71^ Prior work shows that multi-informant approaches improve the accuracy of mental health disorder prediction^60^, and future research should consider incorporating these to improve measurement reliability. Second, our measures of common and rare genetic variation capture only a portion of the total genetic liability to a trait. As genetic association study sample sizes grow and methods continue to advance, the genetic contributions to early-life mental health should be reexamined. Third, our analyses may have been affected by ascertainment bias and non-random missingness across time.^55,72^ To mitigate this, we incorporated weights in MCS and carried out Mendelian imputation of missing parental genotypes in duos with genotype array data. The generally consistent common variant associations observed across cohorts suggest that this mitigation was successful. However, we did not impute missing parental genotypes for the exome sequence data since parents were typically only sequenced if both were available. This reduced our power to detect direct effects of rare variants, and our results may have been influenced by the non-random ascertainment of trios. Lastly, the use of childhood externalising behaviours and cognitive ability assessed at a single time point as mediators may not fully capture the pathways through which these traits may mediate the direct genetic effects of the selected genetic measures on adolescent mental health.

In summary, our findings provide new insights into the genetic architecture of early-life mental health and suggest several promising directions for future research. As larger and more diverse cohorts of sequenced trios become available, it will be important to examine the contribution of genetic variation across the allele frequency spectrum to internalising and externalising problems across development in varied contexts and genetic ancestry groups. Our results also demonstrate that common and rare variants jointly contribute to early-life mental health, highlighting the need to clarify the interplay between these types of genetic variation. Further work in larger cohorts, ideally with more sensitive phenotyping of internalising symptoms at younger ages, is also needed to elucidate direct and indirect genetic effects on internalising traits. Finally, by identifying potential mediating roles for childhood cognitive ability and externalising behaviours, our findings point to early developmental pathways through which genetic risk may unfold, warranting further investigation into the mechanisms that link early traits to later outcomes.

## Methods

### Study design and participants

#### Avon Longitudinal Study of Parents and Children (ALSPAC)

ALSPAC is a prospective birth cohort study investigating genetic and environmental influences on health-related outcomes across the lifespan^56^. The study has been described in detail previously.^55,56,73^ ALSPAC enrolled 14,541 pregnant women in South West England with expected dates of delivery between 1st April 1991 and 31st December 1992. Enrolled children (N=13,988) and their parents were routinely followed up by questionnaires and clinic visits. The study collected biological samples, including DNA, as well as information on health-related measures and environmental exposures. DNA samples were obtained from most children and a subsample of parents. A detailed description of the cohort is provided in the <u>Supplementary Methods</u>. This study uses data collected for 8,074 unrelated children, 5,737 mothers and 1,673 fathers with genetically-inferred European ancestry (See <u>Preparation of</u> <u>Genetic Data: ALSPAC</u>).

The study website (http://www.bristol.ac.uk/alspac/researchers/our-data/) contains details of available data through a fully searchable data dictionary and variable search tool. Ethical approval for the study was obtained from the ALSPAC Ethics and Law Committee and local research ethics committees. Consent for biological samples has been collected in accordance with the Human Tissue Act (2004). Informed consent for the use of data collected via questionnaires and clinics was obtained from participants following the recommendations of the ALSPAC Ethics and Law Committee at the time.

#### Millennium Cohort Study (MCS)

MCS is a prospective, nationally representative, birth cohort following 18,827 children born in the United Kingdom between September 2000 and January 2002.^57,74^ The study employed strategic sampling, intentionally oversampling participants from disadvantaged areas and ethnic minorities in the United Kingdom to maximise the inclusion of groups that are typically under-represented in research.^75^ Seven sweeps of data collection have been conducted since enrollment at ages 9 months and 3, 5, 7, 11, 14, and 17 years. Extensive demographic, health, developmental, and socio economic data has been collected from children and their families.^57^ DNA samples were collected at age 14 from 9,259 cohort members, 8,898 mothers and 5,179 fathers.^76^ This study uses data collected for 5,470 children, 4,541 mothers, and 2,543 fathers of genetically-inferred European ancestry.

MCS has ethical approval from the National Health Service Research Ethics Committee and the National Research Ethics Service.

#### Born in Bradford (BiB)

BiB is a prospective birth cohort that recruited 12,453 pregnant women from Bradford, a city in the north of England, between 2007 and 2011.^58,59^ At the time of recruitment, 41% of the cohort self-reported white British heritage, and 68% of children lived in the most deprived quintile of neighbourhoods in England and Wales. Subsamples of enrolled children have been followed up at various ages for specific substudies and routine health and education data linkage is available for most children. This study uses data collected for 9,041 children and 2,965 of their parents.

Ethics approval for BiB was obtained from the National Health Service Health Research Authority Yorkshire and the Humber (Bradford Leeds) Research Ethics Committee reference: 16/YH/0320 and 16/YH/0062). Mothers were consented to participate in the study upon attendance of a routine antenatal clinic appointment at the city’s primary maternity unit.

### Phenotypic measures

#### Internalising and externalising symptoms

The SDQ was used to assess emotional and behavioural symptoms at several time points in all cohorts. The SDQ is a well-validated, 25-item questionnaire widely used to screen for mental health problems in community samples of children and adolescents.^60^ It may be completed by parents and teachers of children and adolescents and, after the age of 11, by the children themselves. The questionnaire assesses five domains of behaviour: hyperactivity/inattention, conduct problems, peer relationship problems, emotional symptoms, and prosocial behaviour. A score for externalising symptoms is calculated by summing scores for the hyperactivity/inattention and conduct problems subscales, and an internalising symptoms score is calculated as the sum of the peer relationship problems and emotional symptoms subscales^77,78^. This results in a maximum score of 20 for both the externalising and internalising scales, with higher scores reflecting an increased symptom burden.

For the main analyses, parent-reported SDQ scores from ages 5, 7, 11, 14, and 17 in MCS and ages 7, 9, 11, 13, and 16 in ALSPAC were used. The SDQ was completed by mothers in ALSPAC and by the main caregiver in MCS. For uniformity and to minimise potential reporter biases, the analysis was restricted to mother-reported SDQ scores. In BiB, parents completed the SDQ at ages 3, 4, 10, 11, and 12 years. As information on which parent completed the questionnaire was not available for BiB, all parent-reported scores were analysed together.

#### Childhood cognitive ability

Various measures of cognitive ability were collected at different assessment ages in each of the cohorts. For ALSPAC participants, we used performance on the Wechsler Intelligence Scale for Children^79^ at age 8 as a measure of childhood cognitive ability. In MCS, we used a single latent factor derived from various assessments of cognitive ability administered between ages 3 and 7 (**Supplementary Table 14**). The factor, which explained 39% of the variance in cognitive test performance, was derived using the *factanal* function in R and Bartlett method for scoring.^80^

### Preparation of genotype data

Processing and quality control of the genotype array data are described in detail in the <u>Supplementary Methods</u>. Briefly, initial sample-level quality control removed heterozygosity outliers, sex mismatches, and individuals exceeding cohort-specific missingness thresholds (ALSPAC: >3%; MCS: >20%; BiB: >10%). SNP-level filters were applied to exclude variants with high missingness (>1–5%), low MAF (<0.5–1%), or that violated Hardy-Weinberg equilibrium. Multi-batch datasets were merged and KING^81^ was used to remove unexpected relatives, yielding maximal unrelated subsets of genetically inferred European-ancestry children (n≈2,800–8,600 per cohort) and their parents. Genetic ancestry was inferred using principal component analysis and samples with genetically-inferred European ancestry were identified in each cohort. Finally, each cohort was imputed separately (ALSPAC to the TOPMed r2 panel via the TOPMed server, and MCS/BiB to the HRC panel via the Michigan server) and only well-imputed bi-allelic variants (Minimac R² > 0.8 for ALSPAC/MCS; > 0.9 for BiB; MAF > 1%) were retained (≈5.5–8.1 million SNPs per cohort).

### Calculation of polygenic indices

PGIs were calculated using GWAS summary statistics for the following traits: EA^82^, the cognitive and non-cognitive component of EA^83^, ASD^84^, schizophrenia^85^, major depressive disorder^86,87^, generalised anxiety disorder^88^, externalising behaviours^89^, ADHD^90^, neuroticism^91^. SNP weights were calculated using LDpred2-auto^92^, which does not require a validation dataset for tuning of hyper-parameters. LD reference panels were calculated using 1,444,196 HapMap3+ variants^92,93^ and a subset of unrelated European individuals (kinship coefficient < 0.0442) from the target samples, leaving 1,212,346 SNPs for ALSPAC, 1,290,455 SNPs for MCS and 934,933 SNPs for BiB. The --score function in PLINK v1.9 ^94^ was used to calculate the children’s PGIs as the weighted sum of genotypes across a set of SNPs for each individual. Lastly, to adjust for population stratification, the first ten genetic principal components were regressed out of the PGI within each cohort, and the residualised PGI was used in further analysis.

### Mendelian imputation of parental genotypes

For individuals with genotype data available for only one parent, we used *snipar*^14,95^ with default parameters to impute the missing parental genotypes. Parental PGIs were calculated from the observed and imputed parental genotypes (Minimac4 R^2^ > 0.9) using the pgs.py script provided in the *snipar* package. Mendelian imputation increased the number of genotyped trios from 1,471 to 6,799 in ALSPAC, 2,224 to 5,310 in MC, and 313 to 2,466 in BiB. See **Figure 1** for the total number of trios in each cohort post imputation. LDpred2-auto SNP weights (see previous section) were used for parental PGI calculation.

### Exome sequence data preparation

The procedures for generating and quality control of the exome sequencing data are described in detail elsewhere^96^. Briefly, single nucleotide variants (SNVs) and insertions and deletions (indels) were called for samples from ALSPAC (N=11,994), MCS (N=15,050), and BiB (N=11,916) with GATK (version 4.2.4.0 and later releases) following GATK best practices^97^. Sample quality control included the exclusion of suspected sample mismatches, and outlier samples identified using several metrics (e.g., variant counts, heterozygosity). To facilitate variant quality control, a random forest was trained on pre-defined truth sets in each cohort separately. The random forest filtering was then applied with additional genotype-level and missingness filters. Specifically, SNVs were filtered (genotypes were set to missing) using the following thresholds: allele depth (DP) < 5, a heterozygous allele balance ratio (AB) < 0.2, or a genotype quality (GQ) < 20 (ALSPAC) or < 15 (MCS and BiB); Indels were filtered using these thresholds: DP < 10 (ALSPAC and MCS) or < 5 (BiB), AB < 0.3, GQ <10 (ALSPAC) or GQ < 20 (MCS and BiB). Variants that failed the random forest filtering or with missingness > 0.5 after applying the genotype filters were removed. The final dataset included samples for 8,436 children and 3,215 parents in ALSPAC, 7,667 children and 6,925 parents in MCS, and 8,784 and 2,875 children in BiB.

### Rare variant annotation

All variants were annotated using the MANE Select transcript, and loss-of-function confidence (LOFTEE) and deleteriousness scores (CADD) were assigned using the Ensembl Variant Effect Predictor (VEP)^98^. Protein-truncating variants (PTVs) were defined as those classified as “high confidence” by LOFTEE, with a CADD score > 25 (for SNVs), and not located in the last exon or intron. We defined damaging missense variants as those with CADD ≥ 25 and MPC score^99^ ≥ 2. We restricted our analysis to rare variants with a cohort-specific internal allele frequency < 0.1% (allele frequency in a subset of unrelated children) and an external allele frequency in gnom-AD^100^ V3 of < 3x10^-5^. As BiB has two main ancestry groups, European and South Asian, we restricted our analysis to variants with an internal allele frequency < 0.1% in both genetic ancestry groups.^96^

### Calculation of rare variant burden scores

We calculated a rare variant burden score (RVBS) for each consequence class using the following formula, based on work by Gardener et al^39^:

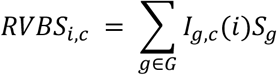

where *I_g,c_*(*i*) is an indicator function for whether an individual i has a variant of consequence c in gene g, *S_g_* is the fitness cost for heterozygous carriers of a pLoF allele in gene i estimated in Sun et al.^61^ , and G is the set of all autosomal genes.

### Non-response and sampling weights for MCS

MCS employed a nonrandom sampling strategy and sampling weights are provided by the study team to adjust for this.^101^ Additionally, we used inverse probability weighting to estimate nonresponse weights to account for the nonrandom missingness of participants at each study sweep. The process of generating the nonresponse weights is described in detail elsewhere.^17^ Briefly, we fit a logistic regression to predict whether an individual had complete data for a given association analysis based on a set of predictors associated with attrition. We then extracted the predicted probabilities of having complete data for all individuals and used their inverse values as nonresponse weights. We incorporated the product of the UK sampling weight and nonresponse weight in our regression models.

### Testing for associations between genetic measures and traits

First, we tested for an association between a genetic measure and externalising and internalising symptoms in ALSPAC and MCS by estimating a linear mixed-effects model using the *lme4* package in R^102^. This analysis was conducted separately in each cohort, restricting to a subset of unrelated participants with genetically-inferred European ancestry. For all regression analyses, genetic measures and age-specific SDQ symptom scores were standardized within each cohort to have a mean of 0 and a standard deviation of 1. Models were estimated for all cohort members and in the subset of cohort members with genetic data (observed or imputed) for the full parent-child trio:

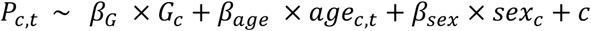

where P is a child’s score for externalising or internalising symptoms at all measured ages regressed on a child’s genetic score G_c_, age at phenotype measurement, and a per-child random effect c. The main genetic effect β_G_ can be interpreted as the average genetic effect on the phenotype across the ages.

To identify outlier associations at specific assessment ages, we also tested for cross-sectional associations at each time point with the following simplified linear model using the *lm* function in R^103^:

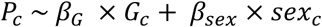

where P_c_ is the symptom score at a single age for child c regressed on the child’s genetic score, age, and sex. The results from these cross-sectional models are shown in **Supplementary Figure 8-10**.

#### Estimating direct genetic effects

Next, we conducted trio-based analyses to assess whether the association between the symptom score and selected genetic measures reflects direct genetic effects or the influence of non-transmitted parental alleles. We estimated regression models adjusting for the parental genetic measures. In these models, the direct genetic effect, δ, may be estimated by subtracting the effects of non-transmitted alleles, θ_NT,_ from transmitted alleles, θ_T,_.^14^ These parameters are estimated in the following model:

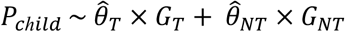

where *G_T_* is the genetic measure for transmitted alleles (the child’s score) and *G_NT_* is the genetic measure for non-transmitted alleles in the parents. This can be further decomposed to:

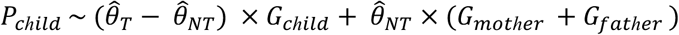

In this model, the coefficient for the child’s genetic measure represents direct effects, and the coefficient on the parent’s genetic measures represents the effects of non-transmitted alleles. As the coefficients on parental genetic measures capture not only indirect genetic effects (sometimes called “genetic nurture”) but also the effects of population stratification and parental assortment^15^, we refer to them as “non-transmitted” rather than “indirect” effects.

In practice, we fitted the following model:

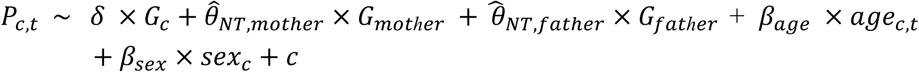

where δ is the direct genetic effect of the child *c*’s alleles, *G_c_*. 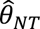 is the effect estimate of the non-transmitted alleles for the mother and father of child *c*.

We also estimated cross-sectional associations using models that control for parental genetic scores:

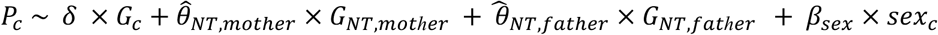

where *P_c_* is symptom measured at a single assessment age and the covariates are the same as above.

#### Testing for age dynamic genetic effects from childhood to adolescence

To test if age impacted the association between a genetic measure and internalising/externalising symptoms, we fitted an additional model with an age-by-genetic score interaction effect:

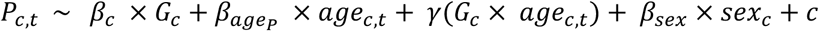

where the main effect β_c_ represents the genetic effect at the first measured age estimated leveraging data at all ages, and *γ* is the change in genetic effect over time.

In trio models, an additional age interaction effect was modeled for the parental genetic values as follows:

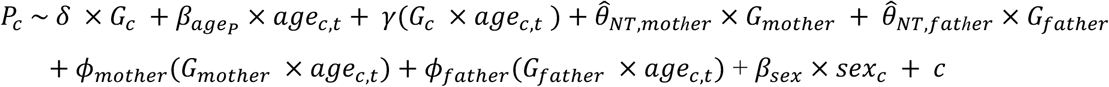

Where *φ_mother_* and *φ_father_* represent the coefficient for the interaction between the parental non-transmitted alleles and child’s age, ie. the change in association with the parental non-transmitted alleles as a function of child’s age.

#### Replication of genetic associations in BiB

We estimated population and direct genetic effects of the common and rare variant measures in BiB using linear models. Due to the modest sample size and low proportion of participants with genetic data and at least two SDQ measurements (<20%), the earliest parent-reported SDQ score for each symptom domain was regressed on each genetic measure and relevant covariates; in trio models, parental polygenic scores were also included as predictors.

#### Correction for multiple testing

As we tested for associations between two correlated outcomes, internalising and externalising symptoms, and multiple correlated genetic measures, we applied the Benjamini-Yekutieli (BY) method to adjust for multiple comparisons, maintaining the false discovery rate (FDR) at 5%. For the main analyses, this correction was applied jointly to all p-values for the main effects of all genetic measures (PGIs and RVBS) from three models: 1) population models including all children, 2) population models restricted to children in trios, and 3) trio models estimating direct genetic effects. Effect size estimates from both MCS and ALSPAC were included in this joint correction. The correction for parental non-transmitted allele effects was applied separately. Lastly, BY correction was applied to association estimates from the BiB models following the same procedure used for the main analyses.

### Stepwise Regression

We used a stepwise regression approach to jointly model common and rare genetic measures that explained variation in internalising and externalising symptoms. For each genetic measure with at a least a nominal association with an outcome in the population models, we ranked the variance explained by the genetic measure by comparing the R^2^ for a baseline model - including age and sex as fixed effects and a per-child random effect - to the R^2^ of a model that included the genetic measure. Models were estimated using a subset of probands with both genotype and exome-sequence data in each cohort (N_ALSPAC_=5,424; N_MCS_=4,500). We then added each genetic measure to the baseline linear mixed effects model in a stepwise manner, entering the genetic measure with the greatest marginal R² first. For each additional genetic measure, we compared the log-likelihood of the model including the new genetic measure to the model without it using the log-likelihood ratio test (LLRT). Predictors were retained if the LLRT was significant (p < 0.05/(number of tests per symptom domain)); p-values were adjusted to account for the number of LLRT per symptom domain in each cohort. We assessed the conditional and marginal R^2^ of the final joint model for each outcome in each cohort separately.

### Mediation Analysis

We tested whether direct genetic effects on adolescent externalising and internalising symptoms were mediated by their effects on two childhood traits, externalising behaviours and cognitive ability. For this analysis, we used scores for the externalising subscale of the SDQ measured at age 5 in MCS and age 7 in ALSPAC as a measure of childhood externalising behaviour. Childhood cognitive ability was defined as described above. Adolescent externalising and internalising symptoms were measured using the SDQ at age 17 in MCS and age 16 in ALSPAC. Analysis was restricted to genetic measures with a nominally significant direct genetic effect on both the mediator and the outcome.

The following structural equation model was fit using the R package *laavan*^104^:

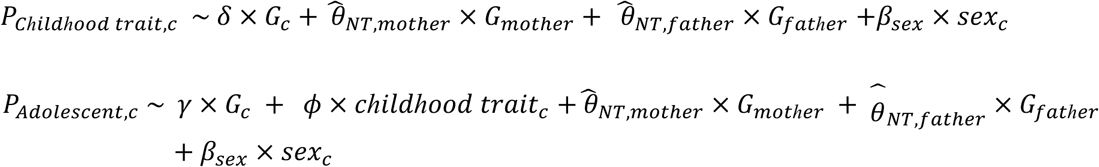

In the first equation, child *c*’s score for childhood externalising behaviour or childhood cognitive ability is regressed on the effect of a child *c*’s genetic measure, G_c_. In the second equation, *ϓ* represents the unmediated effect ofd child *c*’s genetic measure, *G_c_*, and φ represents the effects of the childhood trait on adolescent internalising and externalising symptoms (the indirect pathway). Each model adjusts for sex and the parental genotypes, so δ and *γ* are unbiased by non-transmitted alleles^14^.

The genetic effect on adolescent symptoms mediated by a childhood trait is defined as the product of the effect of the child’s genetic measure on the childhood trait and the effect of childhood externalising symptoms on adolescent symptoms:

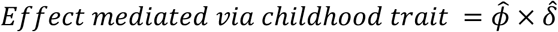

Standard errors were calculated with a sandwich correction to account for non-independence of observation. BY correction was applied to p-values to control the FDR at 5%.

## Data Availability

Researchers can apply to access data from ALSPAC (https://www.bristol.ac.uk/alspac/researchers/access/), MCS (https://cls.ucl.ac.uk/dataaccess-training/data-access/), and BiB (https://bib.vergelabs.dev/research/how-to-access-data/). The ALSPAC study website contains details of all the data that is available through a fully searchable data dictionary and variable search tool: http://www.bristol.ac.uk/alspac/researchers/our-data/.

## Data Availability

https://cls.ucl.ac.uk/dataaccess-training/data-access/

https://bib.vergelabs.dev/research/how-to-access-data/

## Acknowledgements

We thank the Human Genetic Informatics group at the Wellcome Sanger Institute for assisting in the generation and preparation of the whole exome sequencing data for the ALSPAC, BiB, and MCS cohorts and general technical support.

ALSPAC: We are extremely grateful to all the families who took part in this study, the midwives for their help in recruiting them, and the whole ALSPAC team, which includes interviewers, computer and laboratory technicians, clerical workers, research scientists, volunteers, managers, receptionists and nurses.

<u>BiB:</u> Born in Bradford is only possible because of the enthusiasm and commitment of the children and parents in BiB. We are grateful to all the participants, health professionals, schools and researchers who have made Born in Bradford happen.

<u>MCS</u>: The Millennium Cohort Study is only possible due to the commitment and enthusiasm of their participants, their time and contribution is gratefully acknowledged. We are grateful to the Centre for Longitudinal Studies (CLS), UCL Social Research Institute, for the use of these data and to the UK Data Service for making them available. However, neither CLS nor the UK Data Service bear any responsibility for the analysis or interpretation of these data.

## Author Contributions

OW and EW conducted all analyses. OW, DSM, MK, QH, and EW carried out data preparation and quality control with supervision by HCM. MK led the whole exome sequencing data preparation for ALSPAC, MCS and BiB with assistance from DSM and EW. OW, EW, DSM, QH, MEH and HCM provided key intellectual input. HCM supervised the analyses and directed the study. OW wrote the first draft of the manuscript with input from EW, DSM, and HCM. All authors read and commented on the final manuscript.

## Ethics Declaration

All data were collected with informed consent and in accordance with relevant guidelines and regulations. Ethical approval for the ALSPAC was obtained from the ALSPAC Ethics and Law Committee and local research ethics committees. MCS has ethical approval from the National Health Service Research Ethics Committee and the National Research Ethics Service. Ethical approval for BiB was obtained from the National Health Service Health Research Authority Yorkshire and the Humber (Bradford Leeds) Research Ethics Committee reference: 16/YH/0320 and 16/YH/0062).

## Extended Data Figures

**Extended Data Figure 1.**
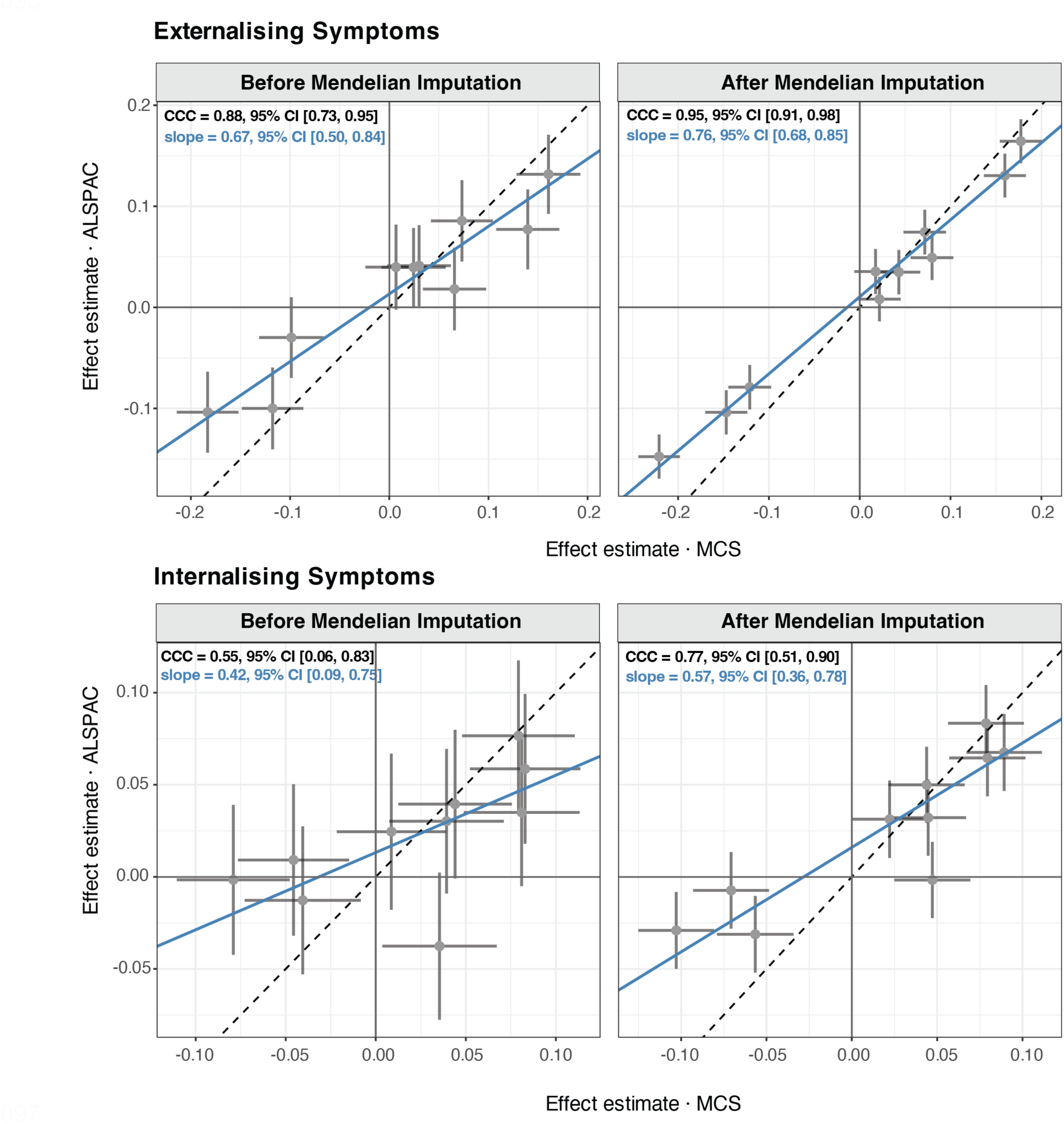
Comparison of population-level associations between ALSPAC and MCS before and after Mendelian imputation of missing parental genotypes. Population-level associations were estimated for children genotyped as complete trios (“Before” Mendelian imputation - both parents genotyped) and children from observed and imputed trios (“After” Mendelian imputation - children genotyped as duos). Points represent standardised effect estimates, and error bars show 95% confidence intervals. Blue line represents the Deming regression fit. Dashed line indicates the y=x axis. CCC is Lin’s concordance correlation coefficient.

**Extended Data Figure 2.**
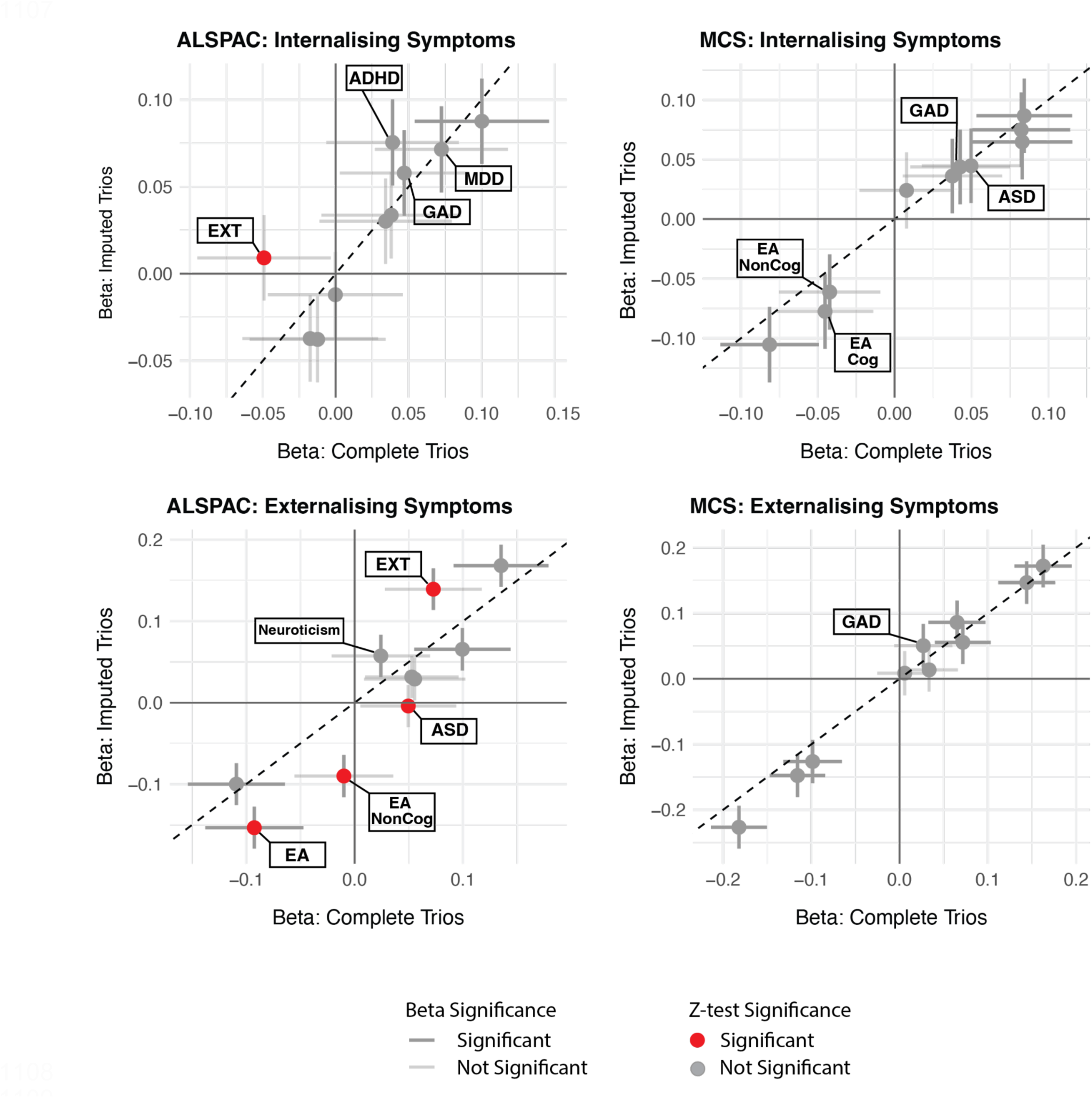
Comparison of standardised effect estimates for population-level associations between polygenic indices (PGIs) and internalising and externalising symptoms using complete versus imputed trios. Population-level associations were estimated for children genotyped as complete trios (both parents genotyped) and children from observed and imputed trios (including children genotyped as duos, the missing parental genotype was imputed using Mendelian imputation). Points represent standardised effect estimates, and error bars show 95% confidence intervals. The opacity of the error bars reflects the significance of the effect estimate. Circles are shaded red where Z-tests indicate a significant difference between estimates derived from complete and imputed trios. Dashed line indicates the y=x axis.

