## Supplementary Information for "The Contribution of Common and Rare Genetic Variation to Emotional and Behavioural Symptoms in Childhood and Adolescence"

### Supplementary Methods

#### Study design and participants

##### *Avon Longitudinal Study of Parents and Children (ALSPAC)*

Pregnant women resident in Avon, UK, with expected dates of delivery between 1st April 1991 and 31st December 1992 were invited to take part in the study. 20,248 pregnancies were identified as being eligible and the initial number of pregnancies enrolled was 14,541.[1–3] Of the initial pregnancies, there was a total of 14,676 foetuses, resulting in 14,062 live births and 13,988 children who were alive at 1 year of age. When the oldest children were approximately 7 years of age, the initial sample was increased by enrolling children who did not join the study originally. The number of new pregnancies not in the initial sample (Phase I enrolment) that are currently represented in the released data and reflecting enrolment status at the age of 24 is 906, resulting in an additional 913 children being enrolled (456, 262 and 195 recruited during Phases II, III and IV respectively). The total sample size for analyses using any data collected after the age of seven is therefore 15,447 pregnancies, resulting in 15,658 foetuses. Of these, 14,901 children were alive at 1 year of age.

Of the original 14,541 pregnancies, 338 were from a woman who had already enrolled with a previous pregnancy, meaning 14,203 unique mothers were initially enrolled in the study. As a result of the additional phases of recruitment, a further 630 women who did not enrol originally have provided data since their child was 7 years of age. This provides a total of 14,833 unique women (G0 mothers) enrolled in ALSPAC as of September 2021.

G0 partners were invited to complete questionnaires by the mothers at the start of the study and they were not formally enrolled at that time. 12,113 G0 partners have been in contact with the study by providing data and/or formally enrolling when this started in 2010. 3,807 G0 partners are currently enrolled.

#### Preparation of genotype data

##### *ALSPAC*

Processing of the genotype array data by the ALSPAC study team was performed in two batches[1] (https://proposals.epi.bristol.ac.uk/alspac_omics_data_catalogue.html#orgc66ee19). Batch one included 8,884 mothers genotyped using the Illumina Human 660W chip and 8,932 children genotyped using the HumanHap550 quad chip. The ALSPAC study team removed samples with missingness rate < 3%, heterozygosity outliers, and mismatched sex as well as SNPs with missingness rate > 5%, MAF < 1%, and HWE test p-value > $1x10^{-7}$. SNPs with missingness rate >1% across all samples in the merged batch were also removed.

Another 2,189 parents (mothers and partners) were genotyped on the CoreExome array chip in the second batch[4]. We applied the following additional quality control filters to the batch two array data: removal of autosomal SNPs with MAF <0.5%, missingness rate >3%, and that failed the HWE test (p-value < $1x10^{-5}$). An additional seven samples with missingness >3% were excluded.

We merged the two batches and used KING[5] for relatedness inference. We excluded 152 samples who had unexpected first-degree relationships but were reported to be from different families. An additional 16 samples that did not match available exome sequencing data supposedly for the same individuals were removed. This left array data for 8,831 children, 9,302 mothers, and 1,706 fathers for further analysis. We identified a maximal subset of unrelated children (N=8,637) using an iterative approach: children with the highest number of genetically inferred relatives (third degree of closer) were iteratively removed until each remaining child had no inferred relatives.

To identify individuals with genetically inferred European ancestry, principal component analysis (PCA) was performed and ALSPAC samples were projected onto samples from 1,000 Genomes phase 3 individuals[6] using the smartpca function from EIGENSOFT version 7.2.1[7]. For this analysis, we used 90,563 (LD)-pruned SNPs (pairwise r2 <0.2 in batches of 50 SNPs with sliding windows of 5) with MAF > 5% after removal of 24 regions with high or long-range LD, including the HLA[8]. All ALSPAC samples project onto European ancestry samples. We then performed PCA on the remaining ALSPAC samples and PC loadings for the first ten principal components were calculated for all participants.

Each batch of genotype array data was imputed separately to the TOPMed r2 reference panel using the TOPMed Imputation Server[9–11]. Variants that had Minimac4 $R^2$ > 0.8 and MAF > 1% in both batches were retained, leaving 8,069,962 variants for downstream analysis.

#### *MCS*

Collection and processing of the MCS genotype array data is described in detail elsewhere[12]. Briefly, MCS participants were genotyped using the Infinium Global Screening Array-24 v1.0. Samples with missingness rate > 20%, high or low heterozygosity (+/- 5 SDs), and sex mismatches were removed. Autosomal SNPs with missing rate <5%, MAF >0.5%, and that passed the HWE test (p-value > $1x10^{-5}$) in a subset of unrelated individuals with genetically inferred European ancestry were retained. An additional 283 samples with missingness rate >5% were removed.

A set of unrelated individuals of genetically inferred European ancestry was identified in MCS following procedures similar to those applied in ALSPAC (as described in Huang *et al.*[13]). Individuals with unexpected familial relationships were removed, leaving 6,153 children, 6,646 mothers and 3,835 fathers.[13]

MCS data was imputed to the Haplotype Reference Consortium[11] (Version r1.1 2016) reference panel using the Michigan Imputation Server[10]. After imputation, multi-allelic SNPs and SNPs with Minimac4 $R^2$ < 0.8 and MAF < 1% were removed, leaving 7,404,321 well-imputed SNPs.

#### *BiB*

BiB samples were genotyped using two chips, the Infinium CoreExome-24 v.1.1 BeadChip (~550 K SNPs), and (2) the Infinium GSA-24 v.1 (~640 K SNPs). Samples with missingness

>10% and sex mismatches were removed. Autosomal SNPs with missing rate <5%, MAF >0.5% and with a HWE test p-value < $1x10^{-5}$ in a subset of genetically-inferred EUR ancestry individuals were excluded.

The two batches were merged and KING was used for relatedness inference. We excluded 465 samples with unexpected familial relationships. Genetic ancestry inference was conducted following the same procedures as in ALSPAC and MCS.[14] Within the subset of genetically-inferred European ancestry children (N=3,019), a maximal set of unrelated children was identified (N=2,832), as described above.

Each batch of genotype array data was imputed to the Haplotype Reference Consortium[11] (Version r1.1 2016) reference panel using the Michigan Imputation Server[10] separately. After imputation, multi-allelic SNPs and SNPs with Minimac4 $R^2$ < 0.9 and MAF < 1% were removed. After merging the batches, 5,462,803 variants remained.

### Comparing main genetic effect estimates between cohorts before and after Mendelian imputation

To compare estimates of the main PGI effects between cohorts before and after Mendelian imputation of missing parental genotypes, we estimated weighted Deming regression models. For each symptom domain, fixed-effect estimates and their standard errors for each genetic measure were extracted from two sets of models: 1) *population effects in trios* and 2) *direct genetic effects* from full trio models. A Deming regression model with the following equation was estimated:

$$\hat{\beta}_{ALSPAC} = \alpha + \beta \, \hat{\beta}_{MCS} + \varepsilon$$

with observation-specific standard errors supplied as weights for both axes.

- Slope ($\beta$) quantifies proportional bias: how closely a one-unit change in MCS corresponds to a one-unit change in ALSPAC.
- Intercept ($\alpha$) quantifies constant bias: whether effect sizes in one cohort are shifted by a fixed amount relative to the other.

Lin's concordance correlation coefficient (CCC) was also computed as a single summary metric that jointly quantifies both constant and proportional bias. Lin's CCC ranges from –1 (perfect discordance) to +1 (perfect concordance).

### Supplementary Figures

Supplementary Figure 1: Overview of externalising and internalising symptom scores across time for each cohort.

Supplementary Figure 2: Comparison of standardised effect estimates for associations between polygenic indices (PGIs) and internalising and externalising symptoms for models estimated using all children versus children in trios.

Supplementary Figure 3: Effect size estimates for parental non-transmitted coefficients (NTCs) for polygenic indices (PGIs) estimated using trio models in MCS and ALSPAC.

Supplementary Figure 4: Associations between polygenic indices (PGIs) and childhood and adolescent externalising and internalising symptoms.

Supplementary Figure 5: Association between PGIs and internalising and externalising symptoms across ages for linear mixed effect models estimated with a PGI:age interaction effect.

Supplementary Figure 6: Effect size estimates for parental non-transmitted coefficients (NTCs) for rare variant burden scores (RVBS) estimated using trio models in MCS and ALSPAC.

Supplementary Figure 7: Associations between rare variant burden scores (RVBS) and childhood and adolescent externalising and internalising symptoms.

Supplementary Figure 8: Cross-sectional associations between polygenic indices (PGIs) and childhood and adolescent externalising symptoms.

Supplementary Figure 9: Cross-sectional associations between polygenic indices (PGIs) and childhood and adolescent internalising symptoms.

Supplementary Figure 10: Cross-sectional associations between rare variant burden scores (RVBS) and childhood and adolescent extermalising and internalising symptoms.

Supplementary Figure 11: Direct genetic effect associations between genetic measures and cognitive ability in MCS and ALSPAC.

Supplementary Figure 12: Comparison of direct genetic effect estimates between ALSPAC and MCS before and after Mendelian imputation of missing parental genotypes.

Supplementary Figure 13: Comparison of standardised direct genetic effect estimates for associations between polygenic indices (PGIs) and internalising and externalising symptoms using complete versus imputed trios.

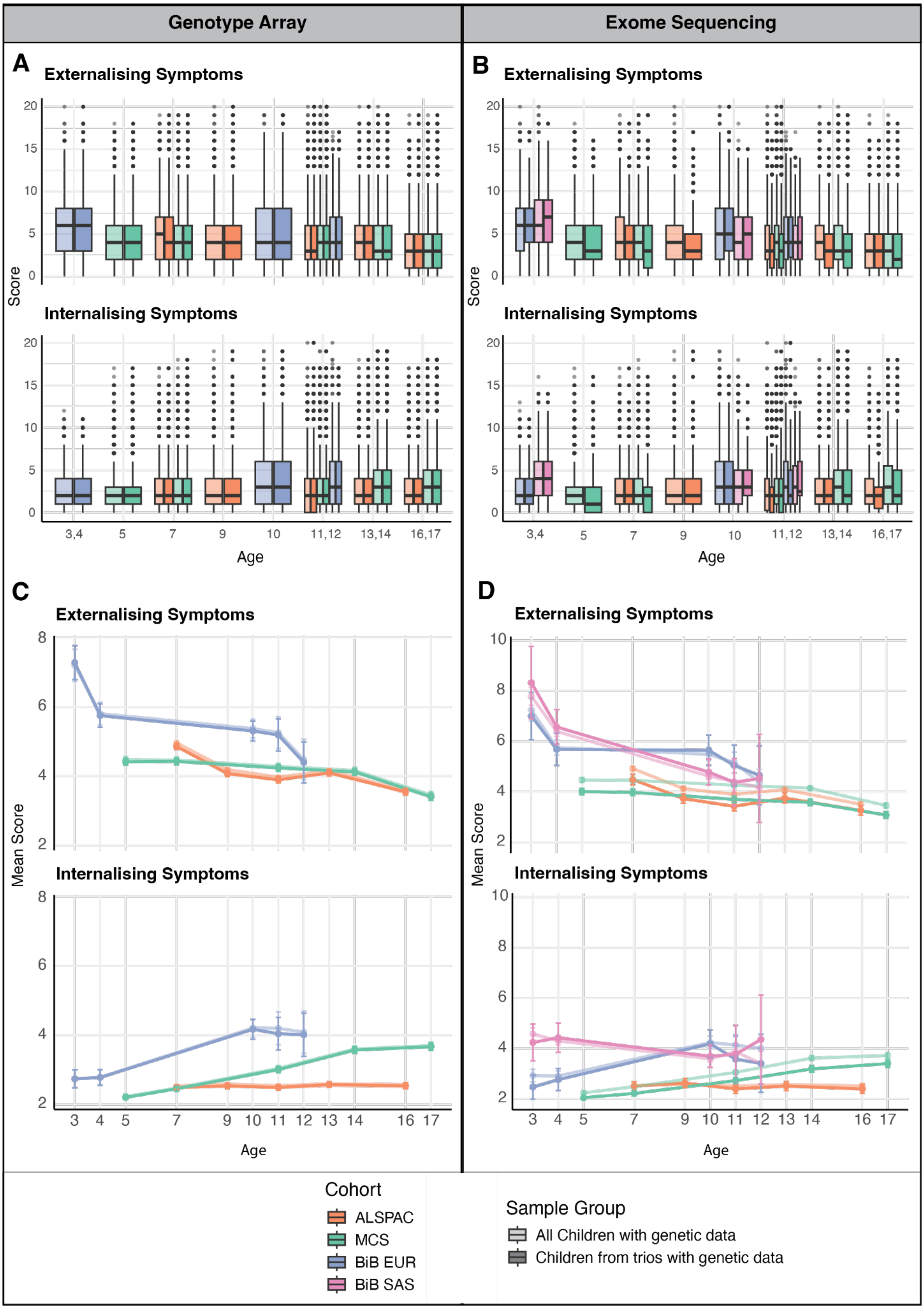


**Supplementary Figure 1. Overview of externalising and internalising symptom scores across time for each cohort. A-B)** Box plots showing externalising and internalising SDQ subscale scores at each assessment age for children with genotype array and exome sequencing data in each cohort (See Figure 1 for sample sizes). Outliers are defined as values more than 1.5 times the interquartile range above the third quartile or below the first quartile. C-D) Mean externalising and internalising SDQ subscale scores at each assessment age for children with genotype array and exome sequencing data in each cohort . Error bars represent 95% confidence intervals. Translucent boxes/lines represent all children with genetic data and opaque boxes/lines represent children from trios with genetic data (including those from imputed trios following Mendelian imputation of genotype array data). ALSPAC: the Avon Longitudinal Study of Parents and Children; BIB: the Born in Bradford study; EUR: genetically-determined European ancestry; MCS: Millenium Cohort Study; SAS: genetically-determined South Asian ancestry.

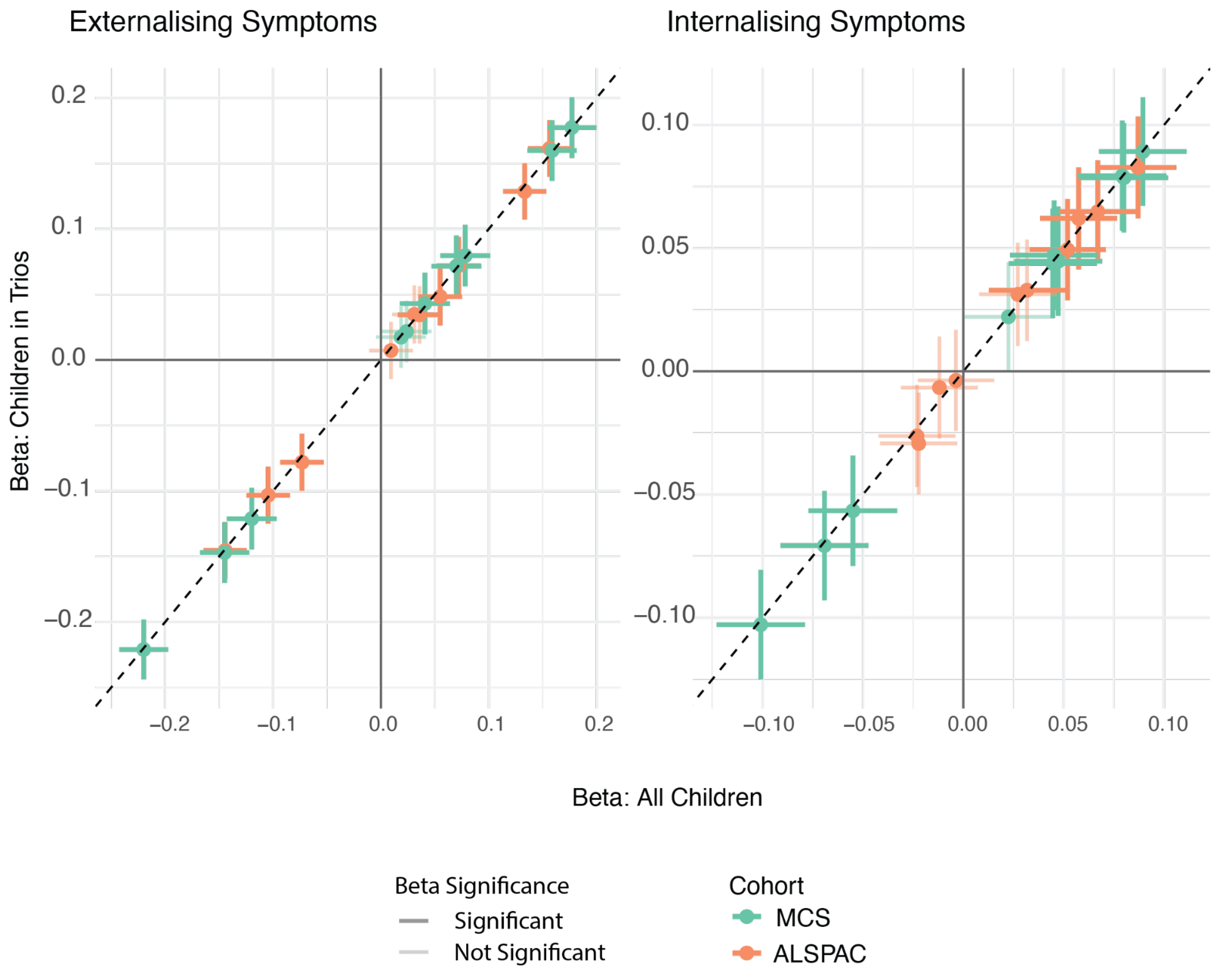


**Supplementary Figure 2. Comparison of standardised effect estimates for associations between polygenic indices (PGIs) and internalising and externalising symptoms for models estimated using all children versus children in trios.** Standardised effect sizes (beta) and 95% confidence intervals are shown for the main effect of each polygenic index, estimated per standard deviation change in symptom score. Children in trios included imputed trios following Mendelian imputation of missing parental genotypes for children genotyped as duos. Dashed line indicates the y=x axis.

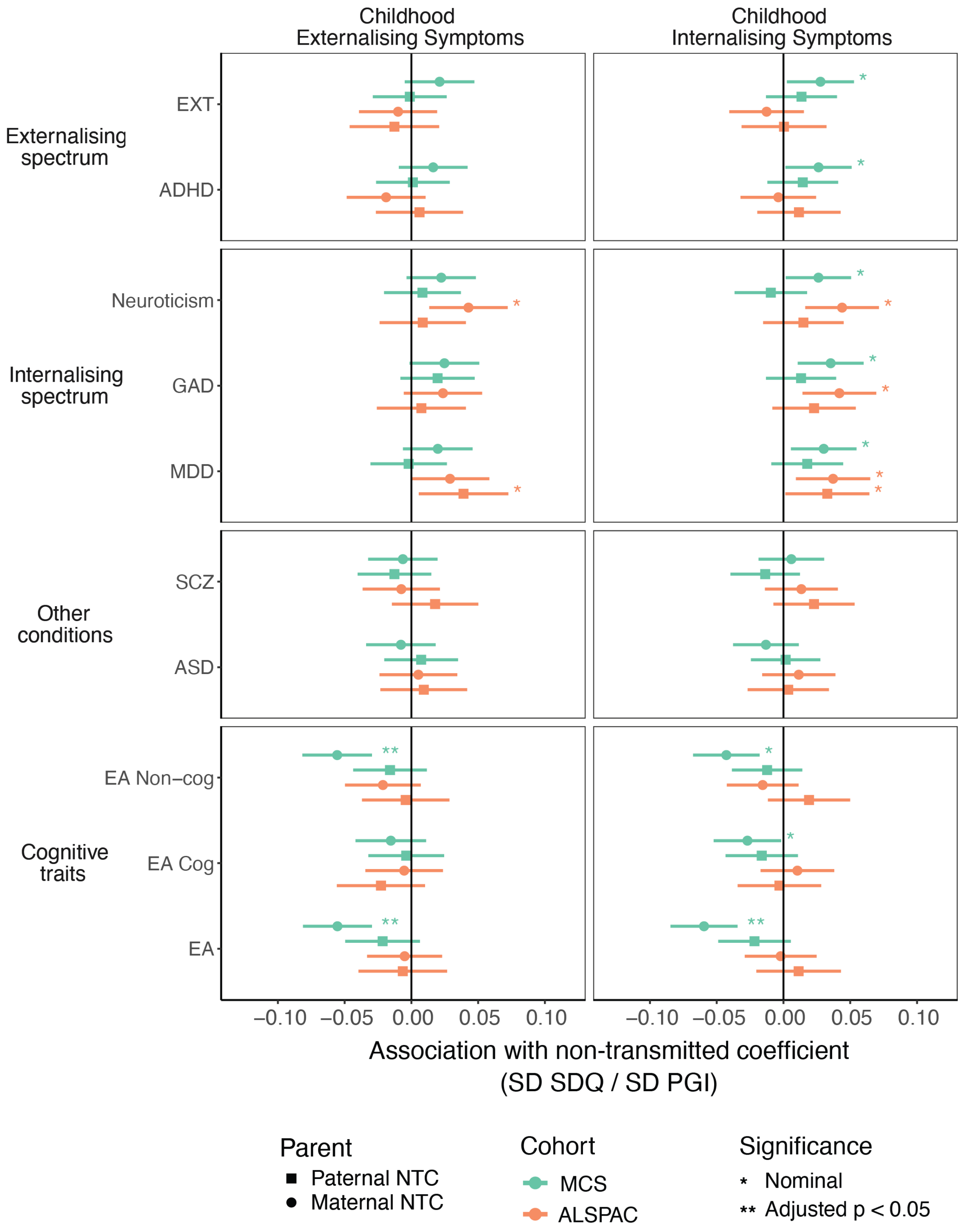


**Supplementary Figure 3. Effect size estimates for parental non-transmitted coefficients (NTCs) for polygenic indices (PGIs) estimated using trio models in MCS and ALSPAC.** Error bars represent the 95% confidence interval for the estimate. ADHD: Attention-deficit hyperactivity disorder; ASD: Autism spectrum disorder; EA: educational attainment; EA Cog: cognitive component of EA; EA Non-cog: non-cognitive component of EA; EXT: externalising behaviours; GAD: Generalised anxiety disorder; MDD: Major depressive disorder; SCZ: schizophrenia.

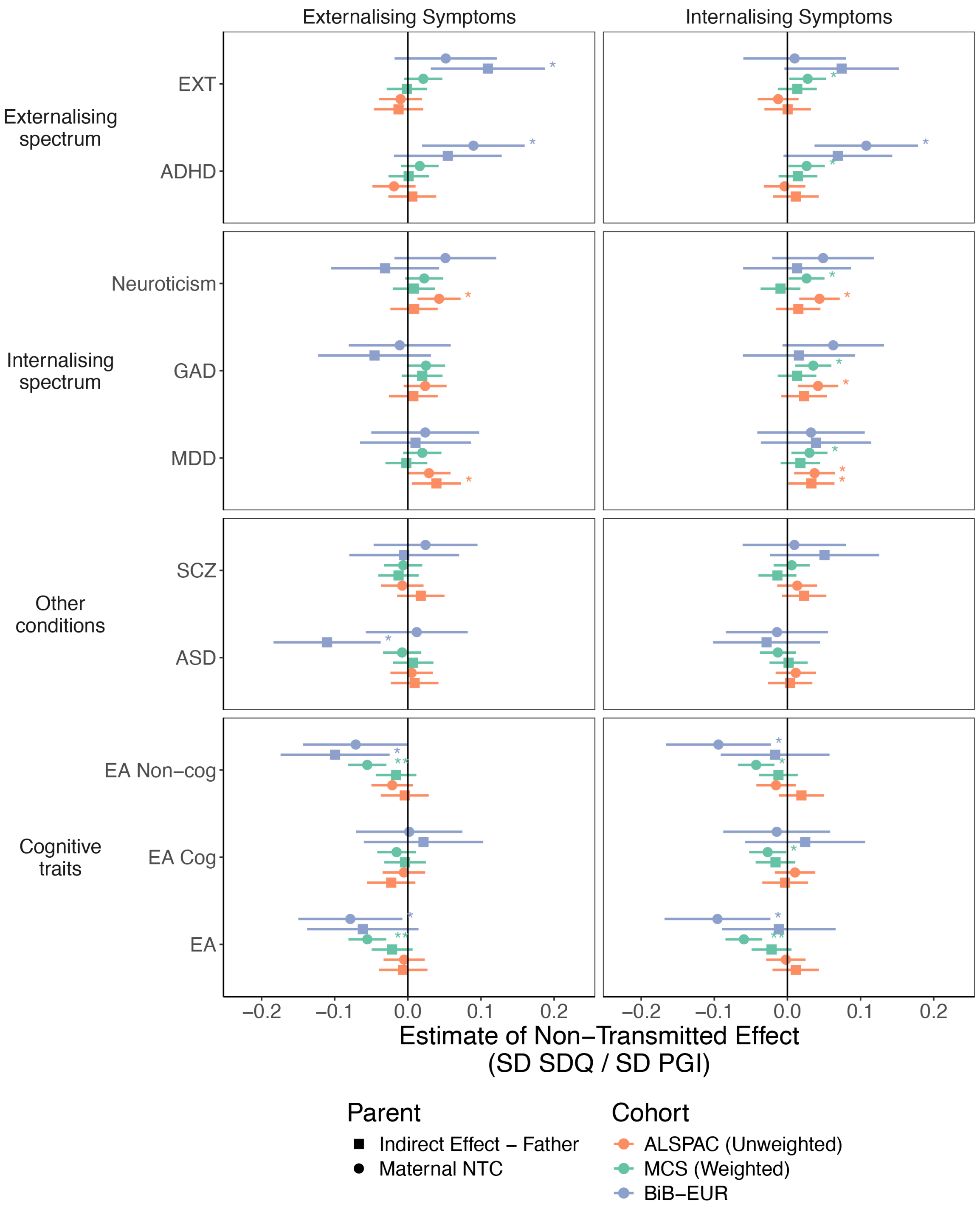


**Supplementary Figure 4. Associations between polygenic indices (PGIs) and childhood and adolescent externalising and internalising symptoms.** Standardised effects and 95% confidence intervals estimated for the main effects of the genetic score per standard deviation change in symptom score. Population effect sizes were estimated in the full sample (triangle) and direct genetic effects were estimated in models controlling for parental genetic scores (shaded circles). Effect size estimates for MCS and ALSPAC were estimated using linear mixed effect models; estimates for BiB were calculated using linear models with the earliest parent-reported SDQ measurement as the outcome. ADHD: Attention-deficit hyperactivity disorder; ASD: Autism spectrum disorder; EA: educational attainment; EA Cog: cognitive component of EA; EA Non-cog: non-cognitive component of EA; EXT: externalising behaviours; GAD: Generalised anxiety disorder; MDD: Major depressive disorder; SCZ: schizophrenia.

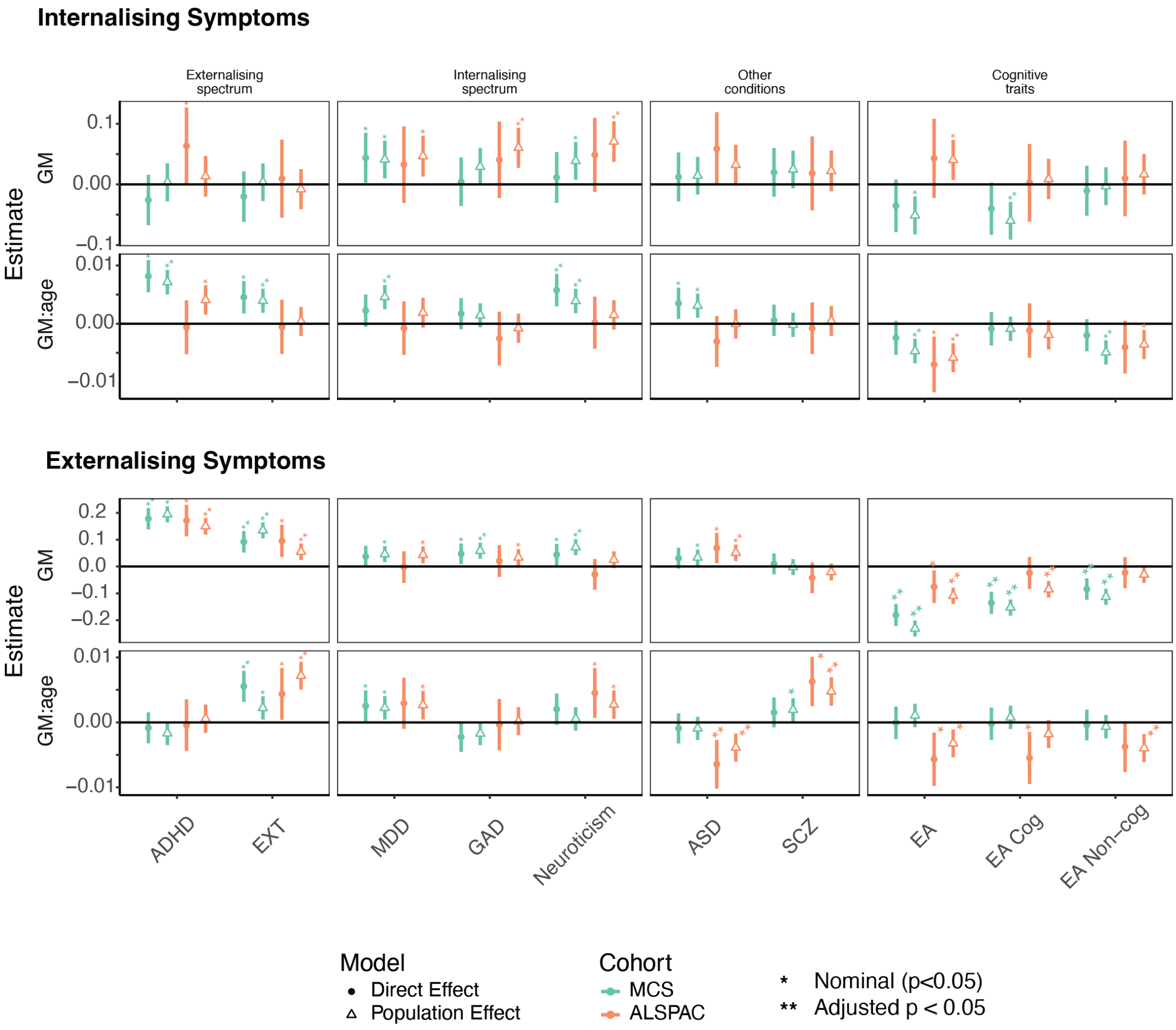


**Supplementary Figure 5. Association between PGIs and internalising and externalising symptoms across ages for linear mixed effect models estimated with a PGI:age interaction effect.** Standardised effect sizes and 95% confidence intervals are shown from linear mixed-effects models including a PGI-by-age interaction term. Estimates are presented for both the main effect of the genetic measure (GM) and the GM × age interaction, per standard deviation change in symptom score. Population effect sizes were estimated in the full sample (triangle) and direct genetic effects were estimated in models controlling for parental genetic scores (shaded circles). ADHD: Attention-deficit hyperactivity disorder; ASD: Autism spectrum disorder; EA: educational attainment; EA Cog: cognitive component of EA; EA Non-cog: non-cognitive component of EA; EXT: externalising behaviours; GAD: Generalised anxiety disorder; MDD: Major depressive disorder; SCZ: schizophrenia.

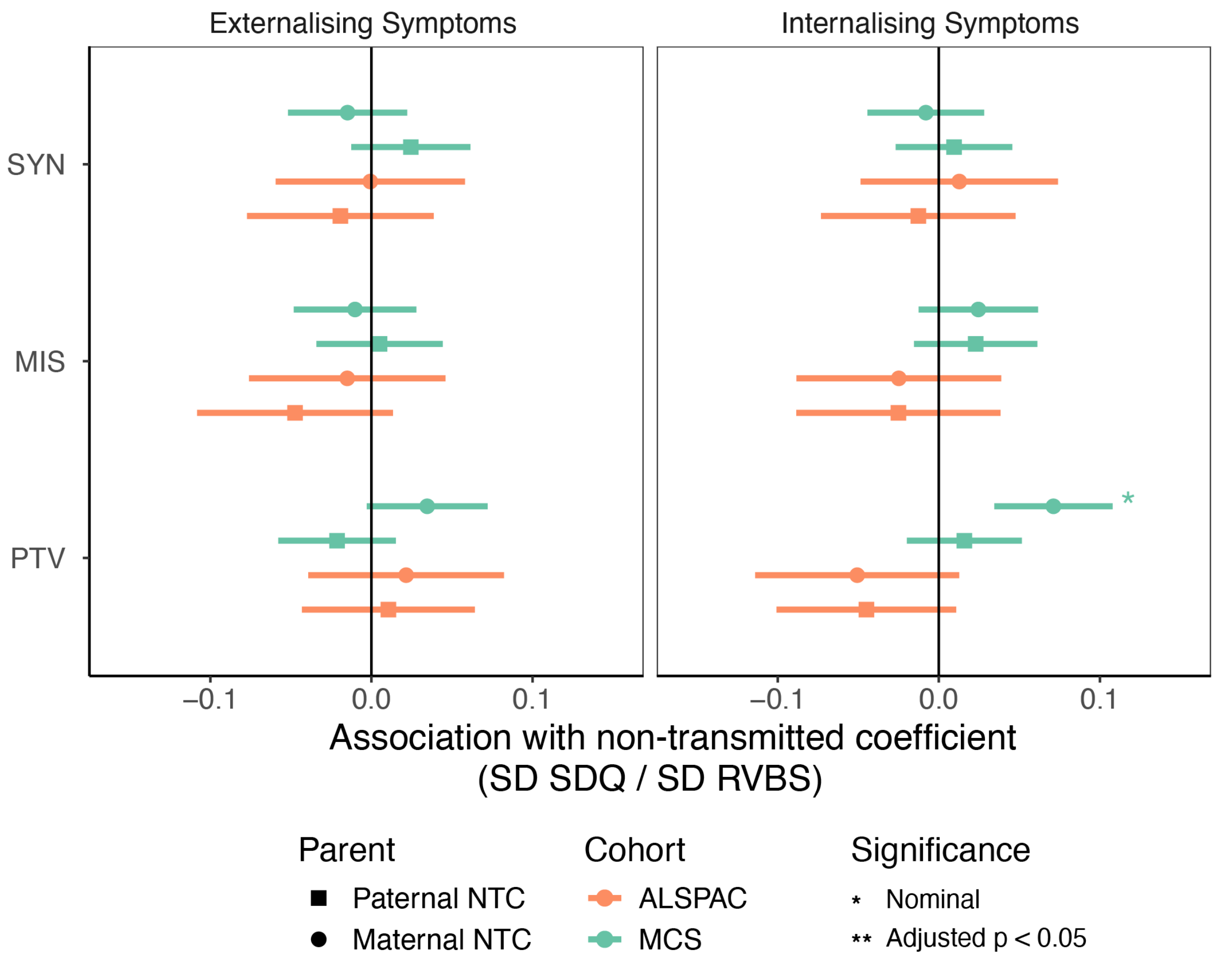


**Supplementary Figure 6. Effect size estimates for parental non-transmitted coefficients (NTCs) for rare variant burden scores (RVBS) estimated using trio models in MCS and ALSPAC.** Error bars represent the 95% confidence interval for the estimate. MIS: missense; PTV: protein truncating variant; SYN: synonymous.

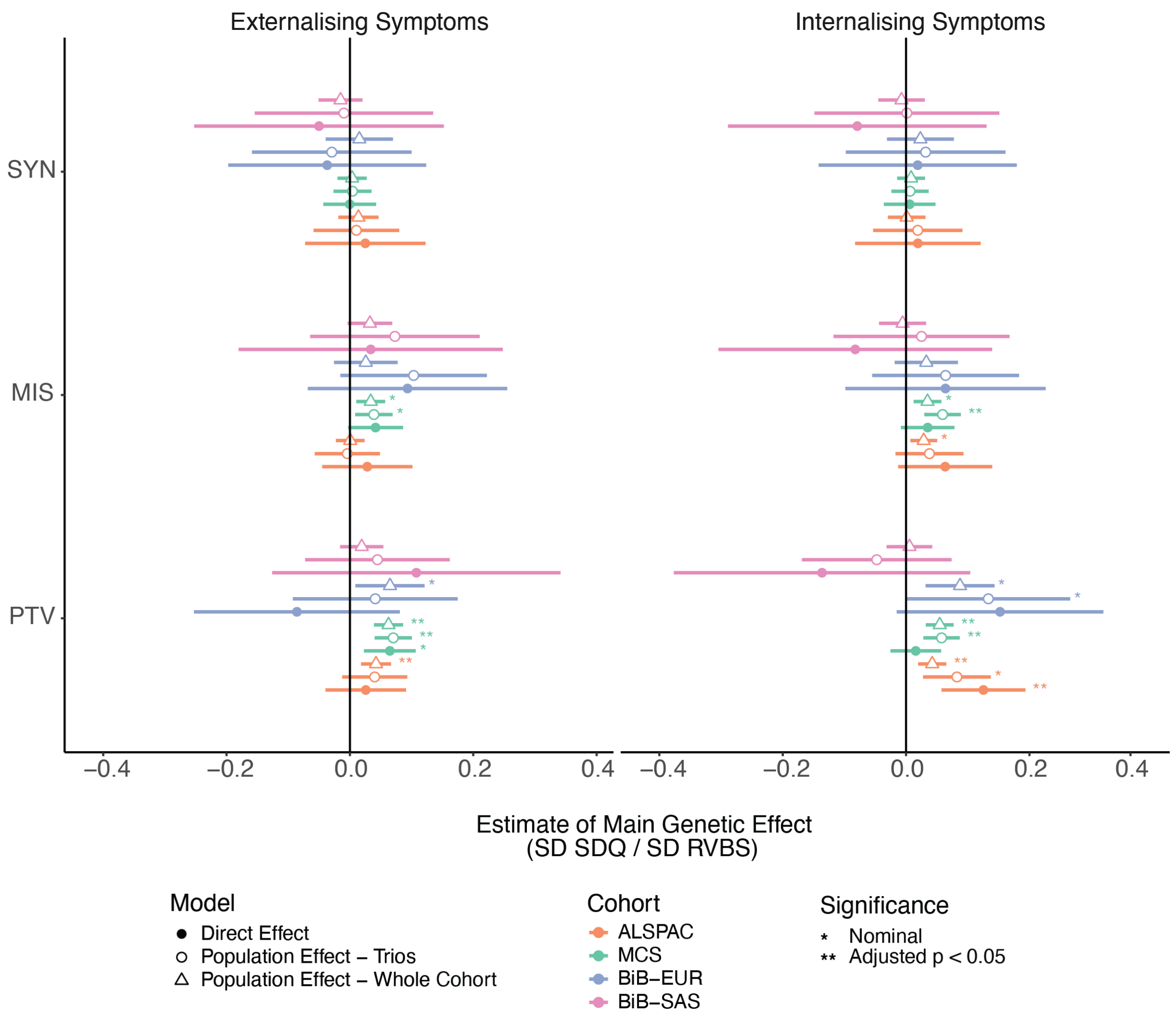


**Supplementary Figure 7. Associations between rare variant burden scores (RVBS) and childhood and adolescent externalising and internalising symptoms.** Standardised effects and 95% confidence intervals estimated for the main effects of the RVBS per standard deviation change in symptom score for RVBS calculated with three different consequence classes. Population effect sizes were estimated in the full sample (circle) and in a subset of children with parental RVBS (triangle). Shaded circles illustrate direct effect estimates from models controlling for parental genetic scores. Effect size estimates for MCS and ALSPAC were estimated using linear mixed effect models; estimates for BiB were calculated using linear models with the earliest parent-reported SDQ measurement as the outcome. MIS: missense; PTV: protein truncating variant; SYN: synonymous.

**Externalising Symptoms**

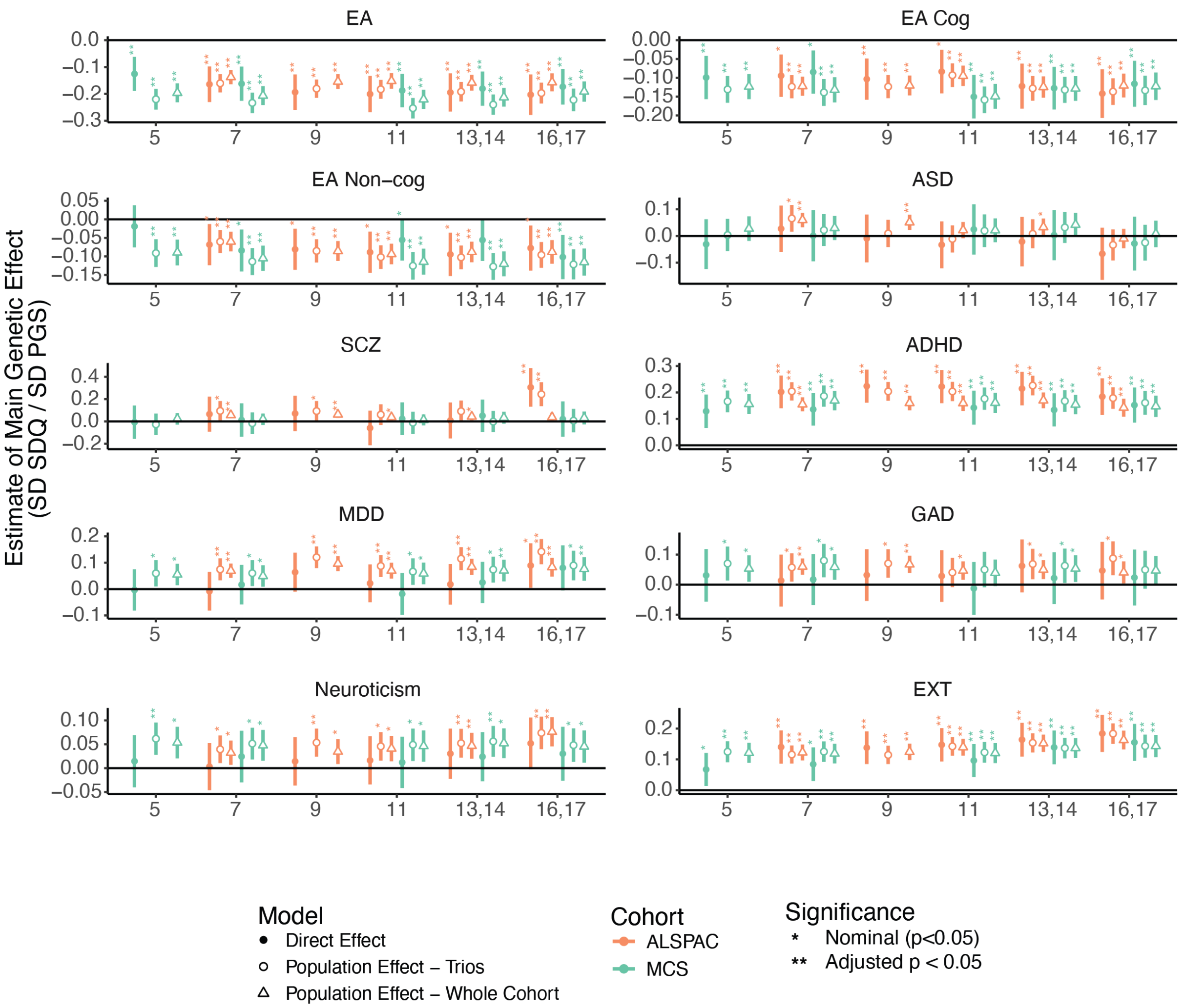


**Supplementary Figure 8. Cross-sectional associations between polygenic indices (PGIs) and childhood and adolescent externalising symptoms.** Effect sizes represent the standard deviation change in SDQ symptom score per standard deviation increase in the PGI, with 95% confidence intervals. Population effect sizes were estimated in the full sample (circle) and in a subset of children with parental PGIs (triangle). Shaded circles illustrate direct effect estimates from models controlling for parental genetic scores. ADHD: Attention-deficit hyperactivity disorder; ASD: Autism spectrum disorder; EA: educational attainment; EA Cog: cognitive component of EA; EA Non-cog: non-cognitive component of EA; EXT: externalising behaviours; GAD: Generalised anxiety disorder; MDD: Major depressive disorder; SCZ: schizophrenia.

**Internalising Symptoms**

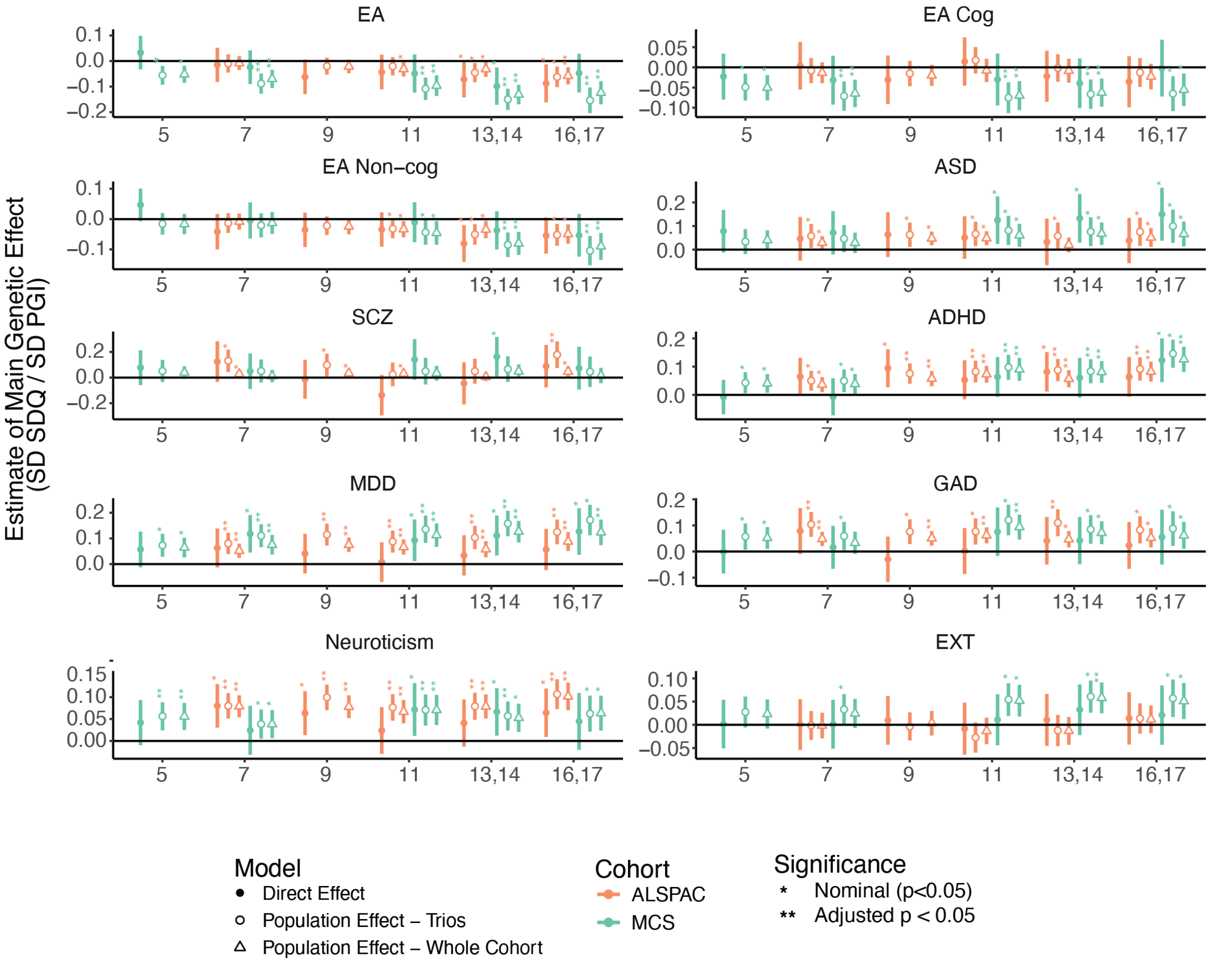


**Supplementary Figure 9. Cross-sectional associations between polygenic indices (PGIs) and childhood and adolescent internalising symptoms.** Effect sizes represent the standard deviation change in SDQ symptom score per standard deviation increase in the PGI, with 95% confidence intervals. Population effect sizes were estimated in the full sample (circle) and in a subset of children with parental PGIs (triangle). Shaded circles illustrate direct effect estimates from models controlling for parental genetic scores. ADHD: Attention-deficit hyperactivity disorder; ASD: Autism spectrum disorder; EA: educational attainment; EA Cog: cognitive component of EA; EA Non-cog: non-cognitive component of EA; EXT: externalising behaviours; GAD: Generalised anxiety disorder; MDD: Major depressive disorder; SCZ: schizophrenia.

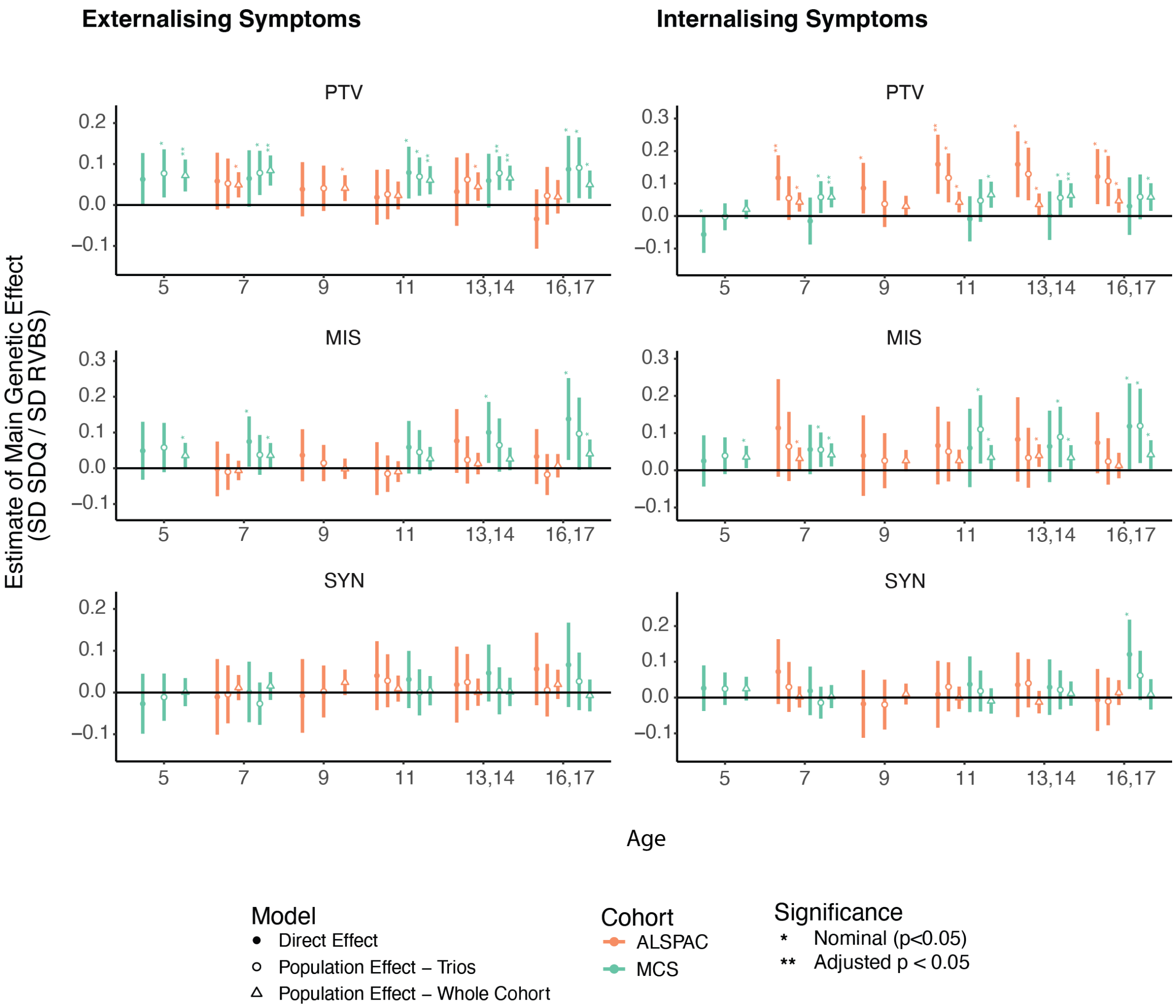


**Supplementary Figure 10. Cross-sectional associations between rare variant burden scores (RVBS) and childhood and adolescent externalising and internalising symptoms.** Effect sizes represent the standard deviation change in SDQ symptom score per standard deviation increase in the RVBS, with 95% confidence intervals. Population effect sizes were estimated in the full sample (circle) and in a subset of children with parental RVBS (triangle). Shaded circles illustrate direct effect estimates from models controlling for parental genetic scores. MIS: missense; PTV: protein truncating variant; SYN: synonymous.

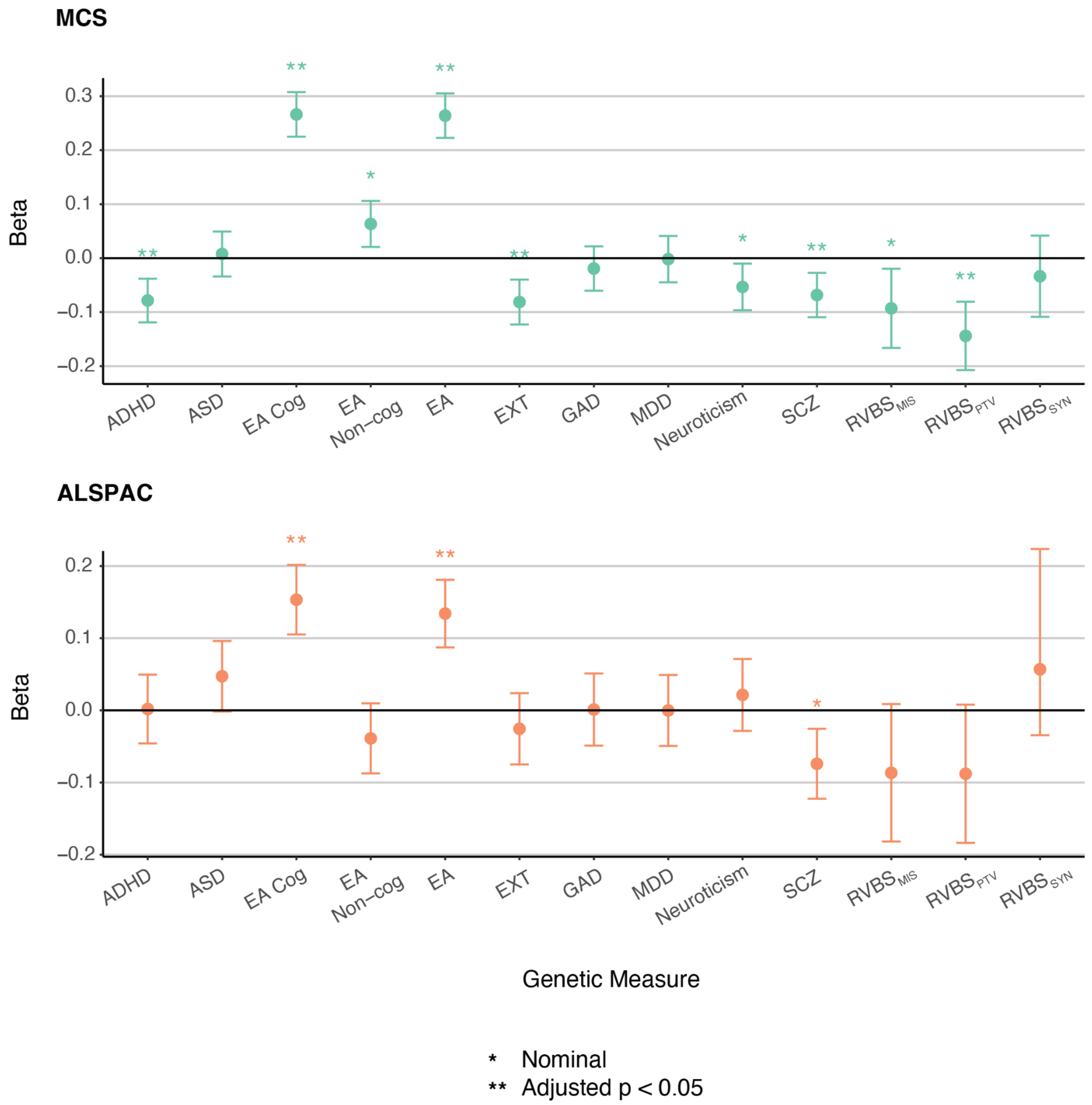


**Supplementary Figure 11. Direct genetic effect associations between genetic measures and cognitive ability in MCS and ALSPAC.** Direct effect estimates are shown from models that control for parental genetic scores, representing the change (in standard deviations) in cognitive ability per one standard deviation increase in the genetic measure. In ALSPAC, cognitive ability is measured as IQ at age 8; in MCS, it is a latent cognitive factor derived from multiple assessments conducted between ages 3 and 7 (see Supplementary Table 13). Error bars represent 95% confidence intervals. ADHD: Attention-deficit hyperactivity disorder; ASD: Autism spectrum disorder; EA: educational attainment; EA Cog: cognitive component of EA; EA Non-cog: non-cognitive component of EA; EXT: externalising behaviours; GAD: Generalised anxiety disorder; MDD: Major depressive disorder; MIS: missense; PGI: polygenic index; PTV: protein truncating variant; RVBS: rare variant burden score; SCZ: schizophrenia; SYN: synonymous.

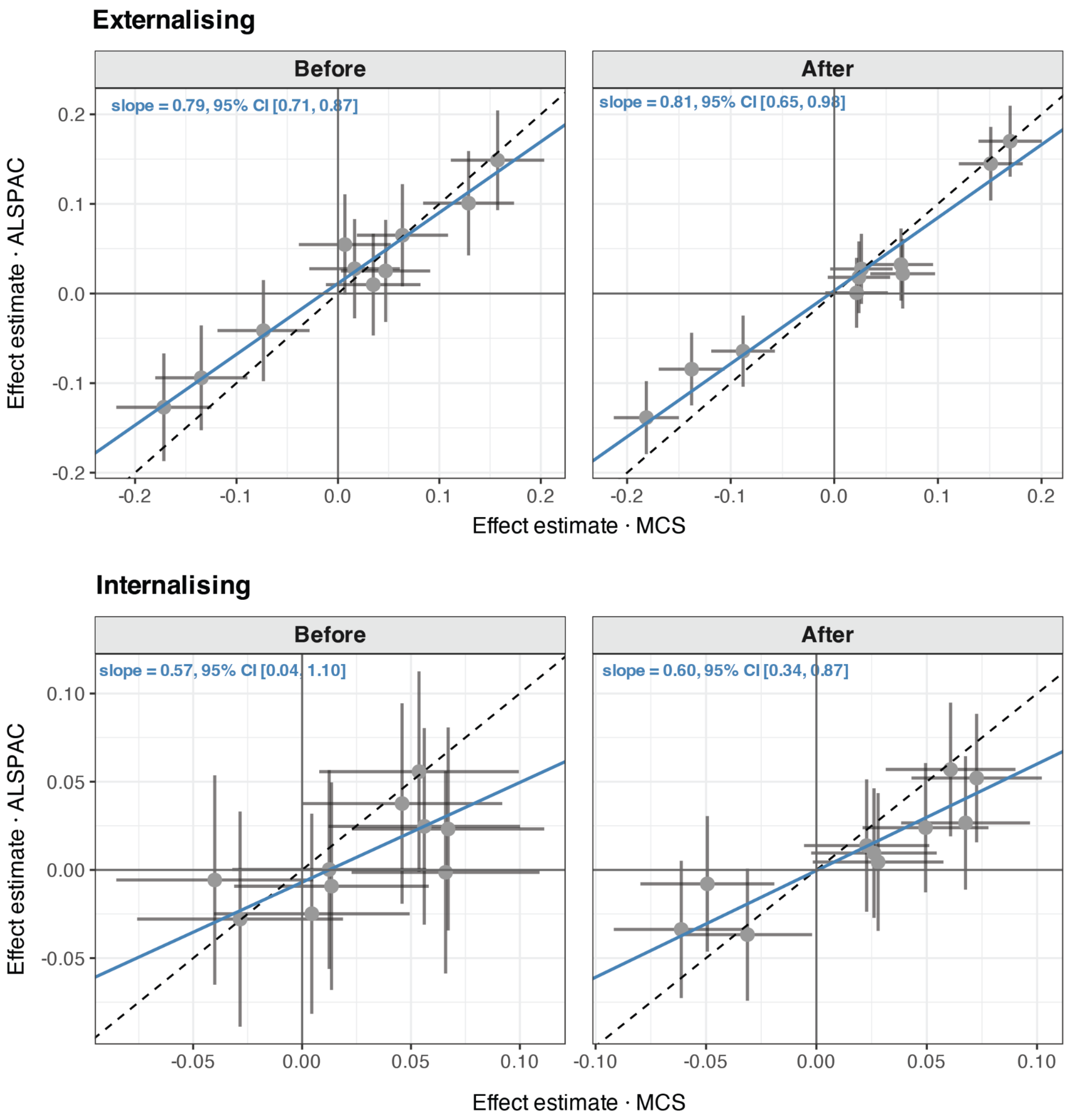


**Supplementary Figure 12. Comparison of direct genetic effect estimates between ALSPAC and MCS before and after Mendelian imputation of missing parental genotypes.** Direct genetic effect associations were estimated using trio models for children genotyped as complete trios ("Before" Mendelian imputation - both parents genotyped) and imputed trios ("After" Mendelian imputation - children genotyped as duos). Points represent standardised effect estimates, and error bars show 95% confidence intervals. Blue line represents the Deming regression fit. Dashed line indicates the y=x axis. CCC is Lin's concordance correlation coefficient.

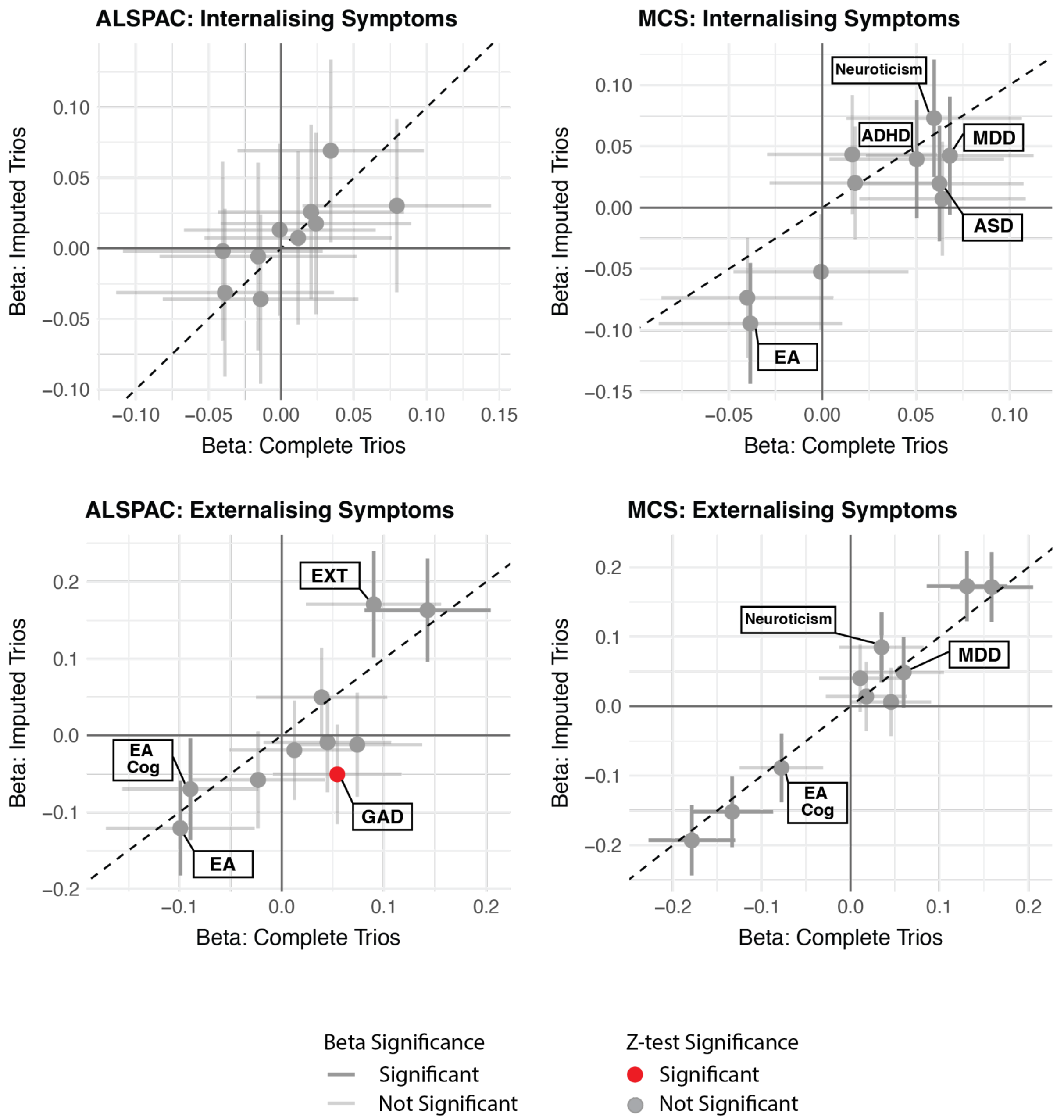


**Supplementary Figure 13. Comparison of standardised direct genetic effect estimates for associations between polygenic indices (PGIs) and internalising and externalising symptoms using complete versus imputed trios.** Direct genetic effect associations were estimated using trio models for children genotyped as complete trios (both parents genotyped) and imputed trios (children genotyped as duos, the missing parental genotype was imputed using Mendelian imputation). Points represent standardised effect estimates, and error bars show 95% confidence intervals. The opacity of the error bars reflects the significance of the effect estimate. Circles are shaded red where Z-tests indicate a significant difference between estimates derived from complete and imputed trios. Dashed line indicates the y=x axis.

### Supplementary Note 1

#### Comparison of main genetic effects between ALSPAC and MCS

To assess the agreement of main PGI effect estimates between cohorts before and after Mendelian imputation of missing parental genotypes, we estimated weighted Deming regression models. These were estimated separately for each symptom domain using population effect estimates from trio-based models (Population Effect - Trios) and direct genetic effect estimates (see Supplementary Methods). Lin's concordance correlation coefficient (CCC) was also calculated to provide a single summary measure of agreement.

For both symptom domains, the Deming regression intercept was not significantly different from zero before imputation, indicating that constant bias was minimal. For externalising symptoms, agreement between ALSPAC and MCS was already high when only observed trios were analysed (Deming slope: population effect 0.67, [95% CI: 0.50-0.84] and direct effect 0.79, [95% CI: 0.71-0.87]) (**Extended Data Figure 1**; **Supplementary Figure 13**; **Supplementary Table 15**). Agreement improved further using imputed genotyped trios (Deming slope: population effect 0.76, [95% CI: 0.68-0.85] and direct effect 0.81, [95% CI: 0.65-0.98]), with the largest improvement observed for population effect models (Lin's CCC: 0.88 before imputation versus 0.95 after imputation) (**Supplementary Table 15**).

For internalising symptoms, agreement was more modest for models estimated using observed genotyped trios only (Deming slope: population effect 0.42 [95 % CI: 0.09–0.75] and direct effect 0.57 [95 % CI: 0.04–1.10]) (**Extended Data Figure 1**; **Supplementary Figure 13**; **Supplementary Table 15**). Imputing missing parental genotypes improved agreement for both population and direct effect estimates (Deming slope: population effect 0.57 [95 % CI: 0.36–0.78] and direct effect 0.60 [95 % CI: 0.34–0.87]), with the largest relative gain seen for population effect estimates (Lin's CCC: 0.56 before imputation versus 0.80 after imputation).

Overall, imputation increased the consistency of PGI effect estimates between cohorts by reducing proportional bias, with the greatest improvements observed for internalising symptoms and population effect estimates.
